# Post-COVID Phenotypic Manifestations are Associated with New-Onset Psychiatric Disease: Findings from the NIH N3C and RECOVER Studies

**DOI:** 10.1101/2022.07.08.22277388

**Authors:** Ben Coleman, Elena Casiraghi, Tiffany J Callahan, Hannah Blau, Lauren Chan, Bryan Laraway, Kevin B. Clark, Yochai Re’em, Ken R. Gersing, Ken Wilkins, Nomi L. Harris, Giorgio Valentini, Melissa A Haendel, Justin Reese, Peter N Robinson, the N3C consortium, the RECOVER Consortium

## Abstract

Acute COVID-19 infection can be followed by diverse clinical manifestations referred to as Post Acute Sequelae of SARS-CoV2 Infection (PASC). Studies have shown an increased risk of being diagnosed with new-onset psychiatric disease following a diagnosis of acute COVID-19. However, it was unclear whether non-psychiatric PASC-associated manifestations (PASC-AMs) are associated with an increased risk of new-onset psychiatric disease following COVID-19.

A retrospective EHR cohort study of 1,603,767 individuals with acute COVID-19 was performed to evaluate whether non-psychiatric PASC-AMs are associated with new-onset psychiatric disease. Data were obtained from the National COVID Cohort Collaborative (N3C), which has EHR data from 65 clinical organizations. EHR codes were mapped to 151 non-psychiatric PASC-AMs recorded 28-120 days following SARS-CoV-2 diagnosis and before diagnosis of new-onset psychiatric disease. Association of newly diagnosed psychiatric disease with age, sex, race, pre-existing comorbidities, and PASC-AMs in seven categories was assessed by logistic regression.

There was a significant association between six categories and newly diagnosed anxiety, mood, and psychotic disorders, with odds ratios highest for cardiovascular (1.35, 1.27-1.42) PASC-AMs. Secondary analysis revealed that the proportions of 95 individual clinical features significantly differed between patients diagnosed with different psychiatric disorders.

Our study provides evidence for association between non-psychiatric PASC-AMs and the incidence of newly diagnosed psychiatric disease. Significant associations were found for features related to multiple organ systems. This information could prove useful in understanding risk stratification for new-onset psychiatric disease following COVID-19. Prospective studies are needed to corroborate these findings.

**Funding:** NCATS U24 TR002306

## INTRODUCTION

Severe acute respiratory syndrome coronavirus 2 (SARS-CoV-2), the virus that causes coronavirus disease 2019 (COVID-19), is responsible for over 600 million cases with 6.5 million deaths worldwide.^1^ Many patients experience manifestations that persist following acute COVID-19 or with onset after the acute period, affecting various organ systems.^2,3^ These manifestations, when not explained by another cause, are part of the broader syndrome of Post Acute Sequelae of SARS-CoV2 Infection (PASC; colloquially referred to as Long COVID). The rate of newly diagnosed psychiatric disease has been found to be significantly increased in patients following COVID-19 infection. The most significant risk has been demonstrated for anxiety disorders, with hazard ratios ranging from 1.3-2.1.^2,4–8^ In a prior study, we found that this risk was only significant in early post-acute phase (28-120 days following COVID-19 diagnosis), reflecting a timeframe when these psychiatric sequelae are most likely to be diagnosed.^6^ A smaller study using electronic health record (EHR) data from a Japanese cohort similarly showed that COVID-19 patients were more likely to receive a diagnosis of psychiatric disease one to three months after COVID-19 compared to controls with influenza or respiratory tract infections.^9^ These findings have major public health ramifications, creating a need to further characterize the risk of newly diagnosed psychiatric disease following COVID-19.

One relevant question is how psychiatric sequelae relate to other manifestations of PASC. Acute COVID-19 may be characterized by neurological manifestations such as confusion, stroke, and neuromuscular disorders. Pathomechanisms include viral neuroinvasion, immune activation and inflammation within the central nervous system (CNS), and endotheliopathy associated with blood–brain barrier dysfunction, which could additionally lead to psychiatric manifestations through diverse pathophysiological mechanisms.^10^ Following acute COVID-19, some patients exhibit manifestations including fatigue, headache, difficulty concentrating, cognitive impairment, anxiety and mood disorders, and dysautonomia.^11^ The pathobiology of these manifestations post-COVID are thought to be the result of a combination of delayed recovery of inflammation, viral persistence, and autoimmunity resulting from infection.^12^ The relationship between newly diagnosed psychiatric sequelae and other PASC manifestations has not been well characterized. One challenge in understanding the pathogenesis of the psychiatric manifestations of PASC is the fact that it is not a single disease; it may have distinct pathogenetic mechanisms underlying different psychiatric manifestations.^13^

In this study, we performed a detailed analysis of 151 PASC-associated manifestations (PASC-AMs) encoded using terms of the Human Phenotype Ontology (HPO), which is widely used to support differential diagnosis and translational research in human genetics.^14^ We analyzed 1,603,767 patients for whom at least 120 days of follow-up data were available after acute COVID-19 in multicenter EHR data from the National COVID Cohort Collaborative (N3C).^15^ We show significant associations between new-onset anxiety, mood, and psychotic disorders and PASC-AMs from six organ systems.

## METHODS

### Study Population and Data Sources

In this retrospective cohort study, we examined the association between PASC-AMs and diagnosis of new-onset anxiety, mood, and psychotic disorders following acute COVID-19. We used patient data accessed through the N3C Data Enclave.^15^ N3C has integrated EHRs from 65 clinical organizations in the United States. We analyzed N3C data frozen on August 31, 2022 which comprised records for 5,858,748 COVID-19 positive patients. Data included over nearly 20 billion rows of data, 1.6 million clinical observations, and 16 million patients. Data were collected from the clinical organizations, normalized to the Observational Medical Outcomes Partnership (OMOP) 5.3.1 vocabulary,^16^ then de-identified and made available to participating N3C research institutions. The study was exempted by the Institutional Review Board (IRB) at the Jackson Laboratory under 45 CFR 46.101(b) (Common Rule). The N3C data transfer to the National Center for Advancing Translational Sciences (NCATS) is performed under a Johns Hopkins University Reliance Protocol # IRB00249128 or individual site agreements with NIH. The N3C Data Enclave is managed under the authority of the NIH; information can be found at https://ncats.nih.gov/n3c/resources.

### Exposures and Outcomes

Clinical data including comorbidities, medications, and outcomes were identified using concept identifiers in the OMOP common data model. Demographics, laboratory values, COVID-19 status, and psychiatric diagnoses were collected for each patient.

Patients were included in the primary analysis if SARS-CoV-2 was detected by polymerase chain reaction (PCR) or antigen test after January 1, 2020. Patients with a history of any primary anxiety, mood, or psychotic disorders prior to 28 days after COVID-19 diagnosis and patients without a medical record covering at least a year prior to and 120 days after COVID-19 diagnosis were excluded from this analysis (Figure 1).

**Figure 1:**
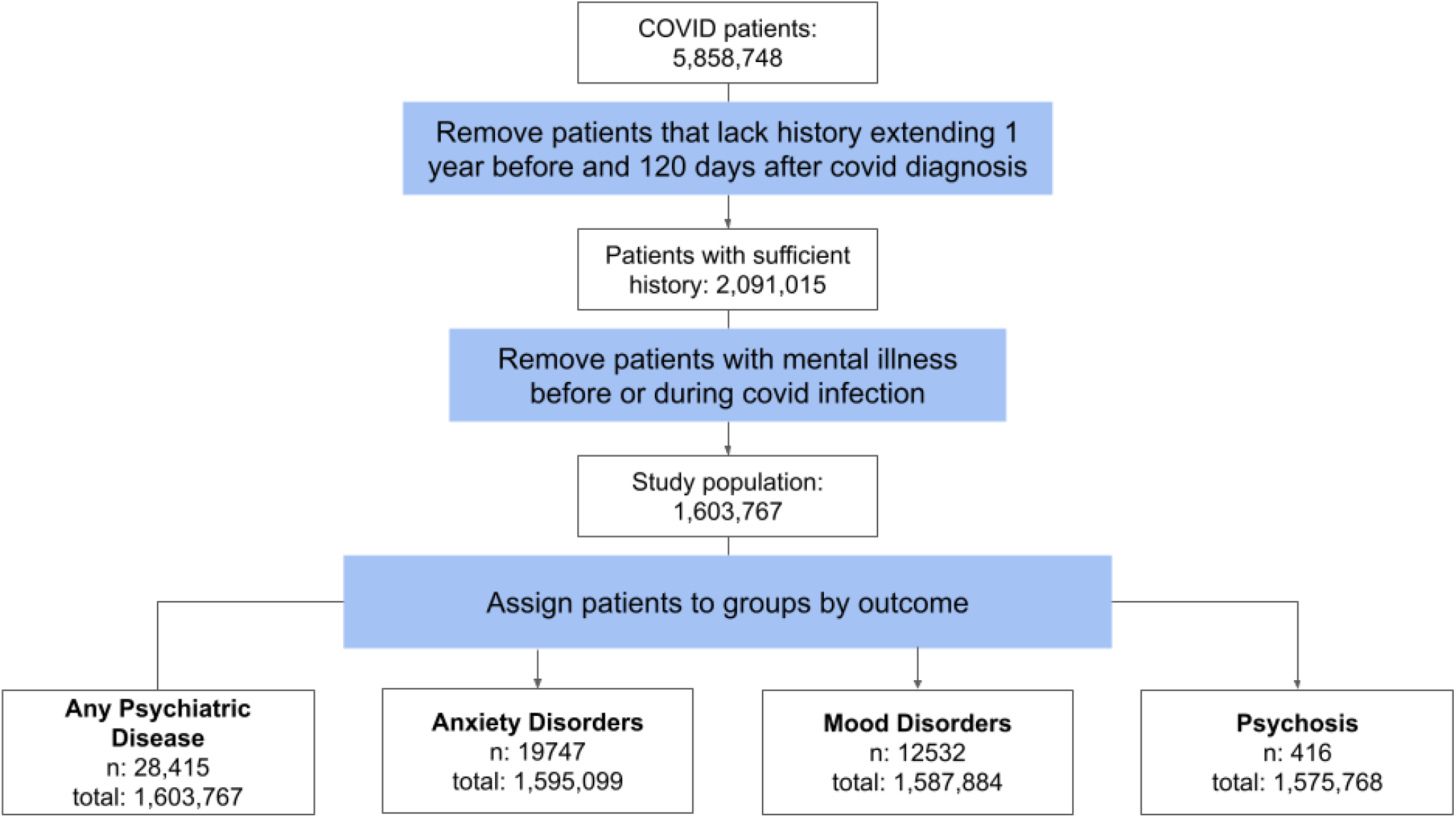
CONSORT-style workflow for creating the cohort. We selected all patients with a positive PCR or antigen test for COVID-19 who had records extending greater than 1 year prior to COVID-19 infection and at least 120 days after diagnosis of COVID-19. Patients with any record of psychiatric disease prior to the post-COVID phase (28 days after diagnosis) were removed. For each outcome, patients with more than one outcome (anxiety, mood disorder, psychosis) whose first psychiatric diagnosis was not the outcome of interest were excluded from the analysis. We report the number of patients included with the outcome (n) and the total number of patients.

We identified patients with a diagnosis of a primary anxiety disorder, mood disorder, or psychotic disorder using diagnostic code defined by OMOP (Supplemental File 1). We treated each category of psychiatric disease as an outcome. Additionally, we performed an analysis with the occurrence of any of the three being retreated as an outcome. Psychiatric outcomes were considered if they were first diagnosed in the period of 28 to 120 days following COVID-19 diagnosis (Figure 2).

**Figure 2:**
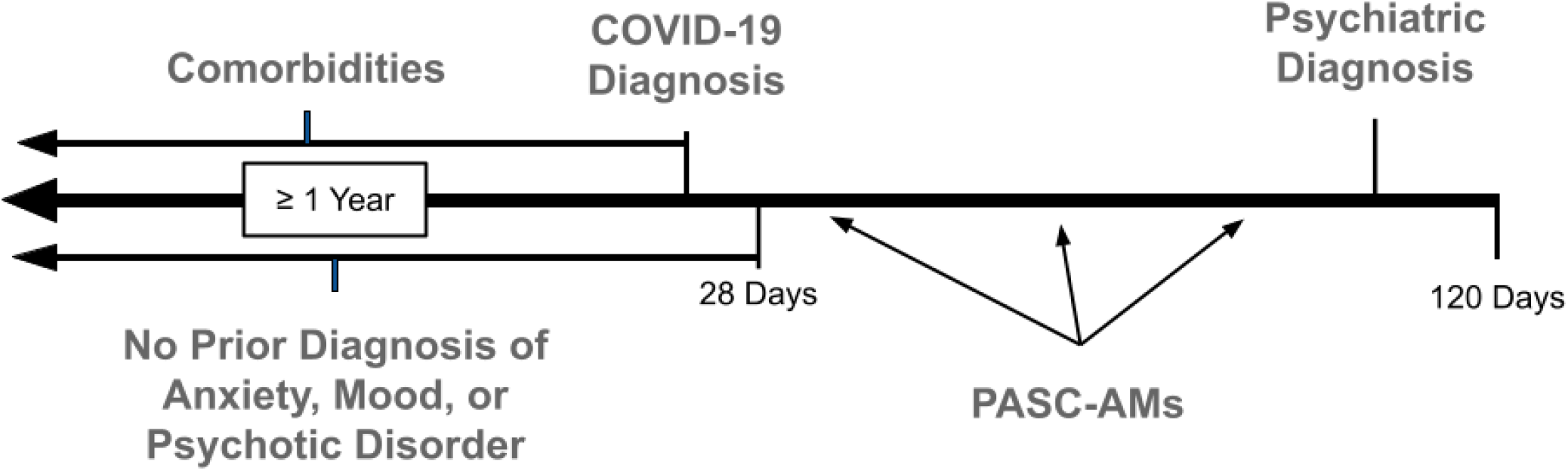
Timeline for measured variables. Every patient was considered with respect to when they were first diagnosed with COVID-19. The pre-COVID phase includes any available records of the patient prior to their COVID-19 diagnosis. Patients whose records did not include at least a year of history prior to COVID-19 diagnosis were removed from the analysis. To focus on patients with a diagnosis of new-onset psychiatric disease, we removed patients with any diagnosis of prior anxiety, mood, or psychotic disorder during their COVID-19 infection (the first 28 days after diagnosis) or anywhere in the pre-COVID phase. Additionally, comorbidities were only considered if they were recorded in the pre-COVID phase. HPO-encoded PASC manifestations were only included if they occured in the early post-COVID phase and prior to the diagnosis of psychiatric disease (if any). PASC-AMs in the early post-acute phase were used to predict risk of psychiatric disease.

We studied known PASC-AMs that had been previously described using 287 Human Phenotype Ontology (HPO) terms.^2,17^ We mapped 176 of these terms to terms in the OMOP data model using the OMOP2OBO mapping algorithm, which leverages well-established automatic mapping techniques and manually-derived expert-verified annotations to align concepts from each source (some HPO terms mapped to multiple OMOP terms and some HPO terms had no good OMOP counterparts).^13,18^ We removed 15 HPO terms from our list of non-psychiatric PASC-AMS because they described psychiatric phenotypes, leaving 161 non-psychiatric HPO terms. 151 of these terms were found in the EHR data and were used for the final analysis. In this way, we were able to identify when patients experienced HPO-defined PASC-AMs using OMOP-encoded patient records. We then categorized HPO terms by affected system. These categories included cardiovascular, constitutional, endocrine, ear-nose-throat (ENT), eye, gastrointestinal, immunology, laboratory, neurological, pulmonary, and skin (Supplemental Tables S1-S11). The HPO terms were recorded in the time period from 28 days after onset of acute COVID-19 up to either the occurrence of newly diagnosed psychiatric disease or 120 days (if no diagnosis of psychiatric disease was recorded) (Figure 2).

In addition to PASC-AMs, we considered the effects of patient demographics, including age, race and ethnicity, sex, smoking status, BMI, visit type (inpatient or outpatient), length of stay (if applicable). We also considered pre-COVID comorbidities (Table 1). All comorbidities were defined as a binary variable indicating whether that patient had received the diagnosis of the comorbidity prior to COVID diagnosis. Only comorbidities present in more than 1% of the population were included in the final analysis (Table 1).

**Table 1.**
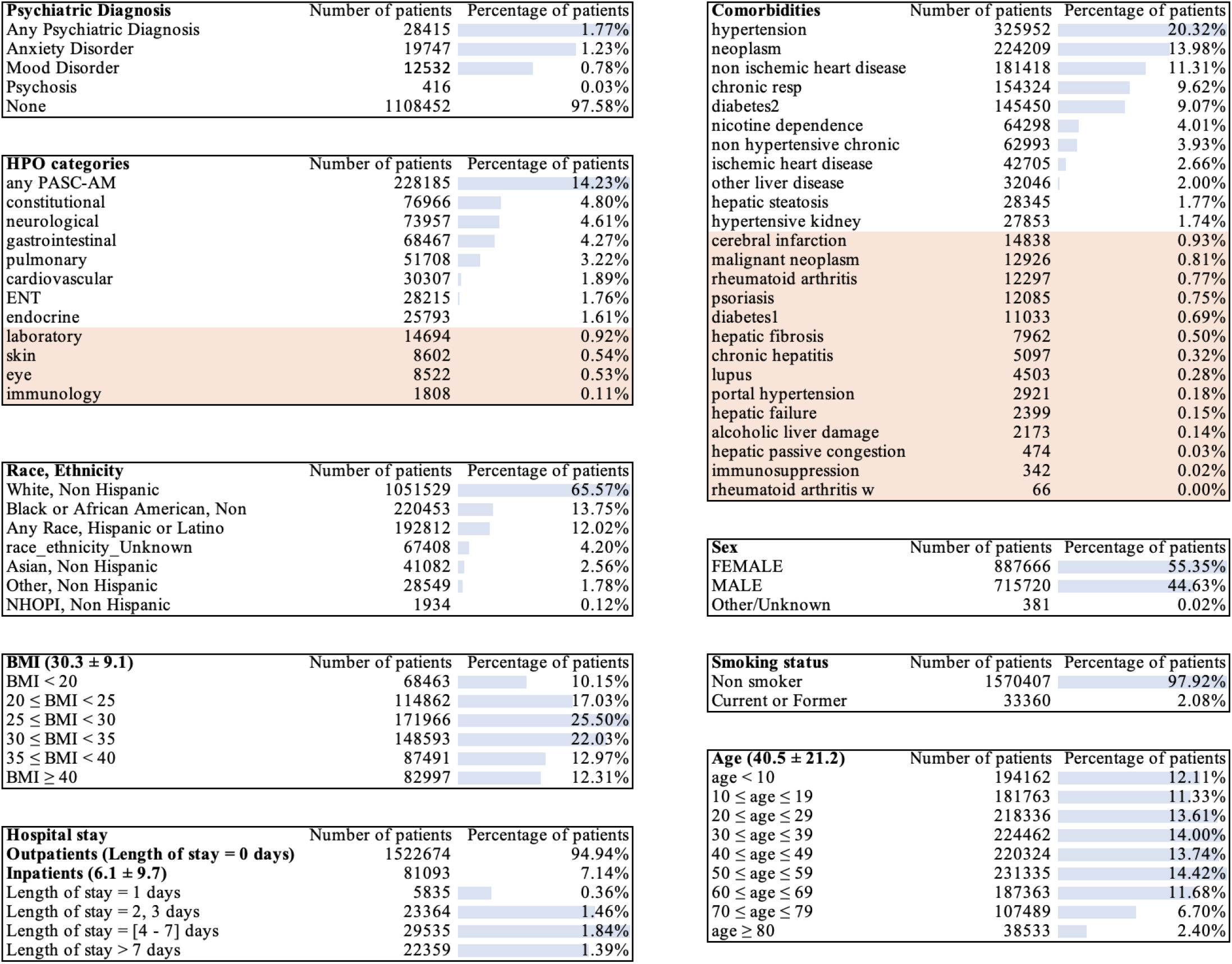
Characteristics of the study cohort are presented as counts (with percentage for categorical/boolean variables) or as mean ± SD (for numeric variables). The percentage is calculated with respect to the size of the entire group (1,603,767). Comorbidities and HPO categories highlighted in orange were removed from the analysis because they were only present in less than 1% of the cohort.

### Statistical analysis

Data analysis was performed using Palantir Foundry (Palantir Technologies Inc., Denver, Colorado). The analysis was structured as a directed acyclic graph of data transformations. Individual transformations were implemented as nodes consisting of SQL, Python, or R code.

To address data missingness, we applied a multiple imputation strategy and computed pooled estimates by applying Rubin’s rule.^19^ BMI was the most commonly missing variable (in 58% of cases). We imputed BMI 23 times with the missRanger algorithm and applied a multiple imputation estimation pipeline to derive the log odds estimates. The missRanger algorithm was chosen based on a previous study comparing different multiple imputation techniques on an N3C cohort of diabetic patients.^20^ We identified all HPO-defined PASC-AMs that occurred between 28 and 120 days following COVID diagnosis. PASC-AMs that were first documented after or on the same day as the diagnosis of a psychiatric disease were not included in the analysis. For each patient, we counted the number of unique HPO terms in each of the eleven categories described in the preceding section. PASC-AM categories present in less than 1% of the population were not considered.

To investigate the association of PASC manifestations and other covariates with newly diagnosed psychiatric disease, we performed logistic regression using the *glm* function in R. The predictors included age, sex, race, pre-existing comorbidities, and the counts of manifestations in each of the eleven HPO categories. Only predictors present in more than 1% of the cohort were used. A patient was said to have the outcome of a newly diagnosed psychiatric disease when it was the first diagnosis of any anxiety, mood, or psychotic disorder seen in the patient’s record. We conducted separate analyses to predict the occurrence of three categories of psychiatric disease: anxiety, mood disorders, and psychosis. The control group was patients with no diagnosis of anxiety, mood disorders, or psychosis. Logistic regression was also applied to predict the diagnosis of any new psychiatric disorder regardless of category. For each regression, we recorded the estimated odds ratio, 95% confidence intervals, and corresponding p-value.

To better understand the temporal relationship between PASC-AMs and psychiatric diseases, we looked at the distribution of patients by time between PASC-AM and outcome. Non-exclusive groups were created for each PASC-AM category that contained any patient with the respective PASC-AM and any psychiatric outcome. Histograms were created to show the distribution across time (ranging 1-92 days) for patients in each group.

We performed a chi-squared test of independence to assess if the incidence of HPO terms in each category were significantly associated with an outcome. For each term, a contingency table was constructed containing the counts of study participants with or without the corresponding HPO annotation and with or without the corresponding new-onset psychiatric disease. For the chi-squared analysis, we removed 3,107 patients (11.3% of those with a psychiatric diagnosis) who had more than one outcome. P-values were adjusted using Bonferroni correction.

### Sensitivity Analyses

Imputation was used to compensate for significant missingness of BMI data. Since BMI is associated with risk of severe COVID-19^21^ and could therefore act as a confounder, we also performed the analysis without imputation, keeping only patients with complete data. To ascertain whether our results are consistent between inpatients and outpatients, we separated our cohorts into those who were admitted for their COVID-19 infection and those who were not.

Many studies have investigated PASC and related manifestations. These studies have used different criteria to identify whether a manifestation is attributed to acute COVID-19 or PASC. To determine whether this had a large effect on our results, we repeated our regression analysis using the alternative definition of 42 days after initial diagnosis for outpatients and 365 days after discharge for inpatients.

In this study we investigate the association between PASC-AMs and subsequent psychiatric disease. In our main analysis we assume that psychiatric diseases diagnosed after PASC-AMs likewise have an onset subsequent to the date on which PASC-AMs were recorded. We performed an additional sensitivity analysis to assess whether our results could have been impacted by a greater delay between onset of psychiatric disease and diagnosis and recording of the diagnosis in the EHR. For this, we excluded PASC-AMs that were diagnosed within ten days prior to the psychiatric diagnosis, and otherwise repeated the analysis unchanged.

## Results

A total of 5,858,748 patients with prior COVID-19 were assessed. We restricted analysis to patients with no previous recorded psychiatric illness and at least one year of data prior to acute COVID-19 diagnosis and 120 days after. This left 1,603,767 patients in the COVID-19 cohort. We found that 1.77% of patients had a newly diagnosed anxiety, mood, or psychotic disorder in the early post-acute phase (28-120 days after COVID-19 diagnosis) (Table 1).

The outcomes of interest were psychiatric diagnoses: anxiety disorder, mood disorder, psychosis, and the combination of any of these three. To identify risk factors we exploited the hierarchical structure of the HPO to group into eleven categories the HPO terms describing clinical manifestations that may be observed in PASC.^2^ We refer to these manifestations as PASC-AMs to reflect the difficulty of inferring causal relationships on the basis of EHR data. The patient’s symptomatology in each category is summarized by an integer-valued variable equal to the number of distinct HPO terms recorded in that category. Constitutional and neurologic PASC-AMs were most common, occuring in 4.80% and 4.61% of patients respectively. A logistic regression was performed with these variables as well as binary variables derived from 25 pre-existing comorbidities and age, sex, and race. We then removed categories and variables which were recorded in less than 1% of the cohort (Figure 2).

We found that there was a significant positive association for four of the seven investigated HPO categories with newly diagnosed psychiatric disease, with the estimated odds ratio ranging from 1.13 to 1.35 for the four categories (Figure 3). Conversely, endocrine PASC-AMs were negatively associated with psychiatric diagnoses with an odds ratio of 0.84 (0.77-0.92, 95% CI). When looking at anxiety disorders alone, all significant associations were retained. Additionally, there was a significant association between ENT symptoms and anxiety disorders. Of the seven HPO categories, three were significantly positively associated with newly diagnosed mood disorder. The strongest single association was for neurological features, with an odds ratio of 1.27 (1.21-1.34 95%CI). Endocrine PASC-AMs had a significant negative association with new-onset mood disorders. Fewer patients were available with newly diagnosed psychosis, but there was still a significant increase in risk for patients with neurological and cardiovascular manifestations for being diagnosed with psychosis (Figure 3). Similar results were found for the sensitivity analyses of stratifying inpatients and outpatients, removing patients with any BMI missingness, and varying the definition for the early post-acute phase (Supplement Tables S13-16). Finally, we performed a sensitivity analysis in which PASC-AMs that were recorded within ten days prior to the new-onset psychiatric disease were removed. We found similar results although a few categories were no longer significant and a negative association between ENT PASC-AMs and mood disorders gained significance (Supplement Table S17).

**Figure 3:**
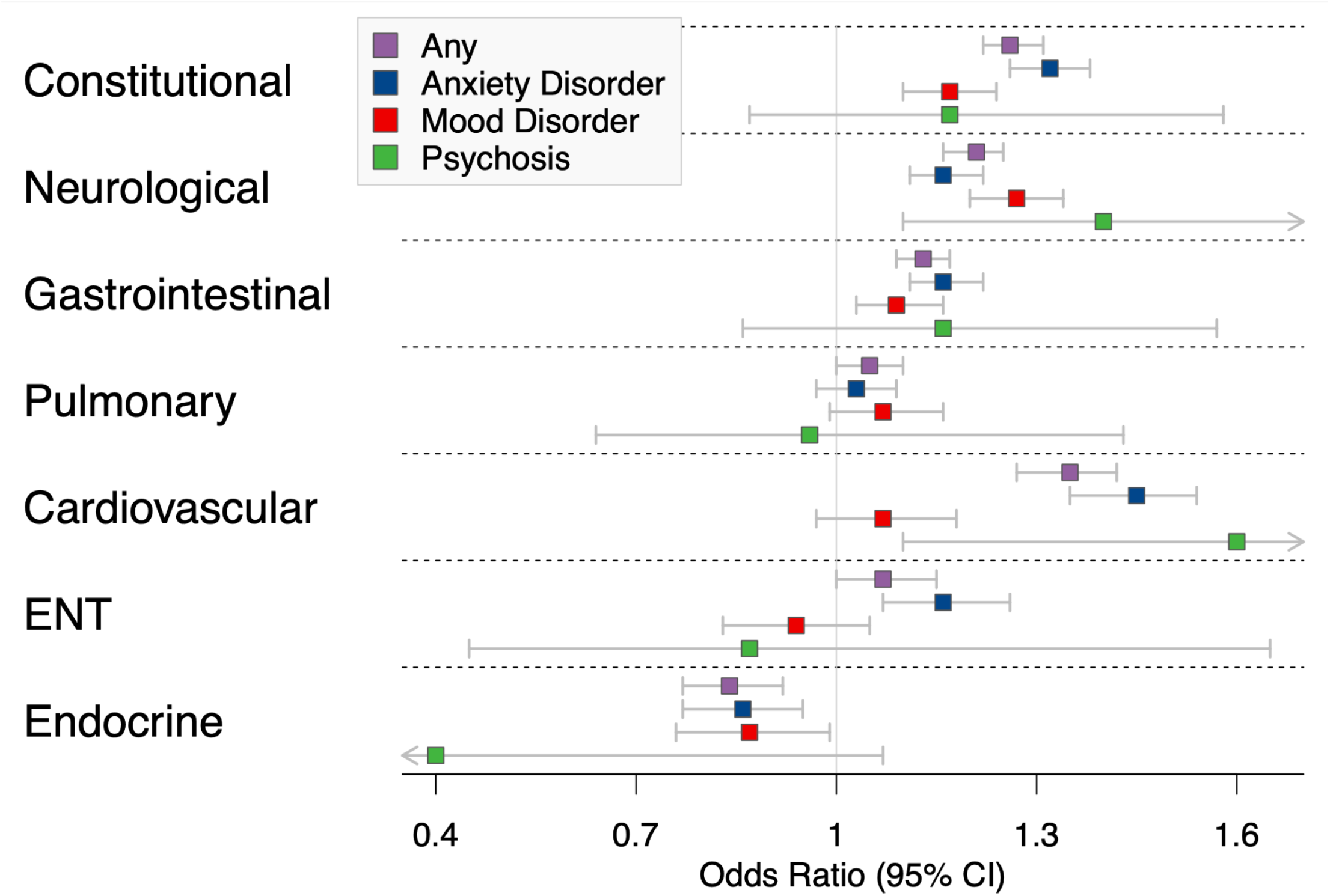
Association of Manifestations to Any Mental Disorder. Odds ratios and 95% confidence intervals for the association of features in the seven investigated HPO categories with all newly diagnosed psychiatric disease and the subcategories anxiety disorder, mood disorder, dementia, and psychosis. See also Supplemental Table S13 for complete results including comorbidities and demographics.

**Figure 4:**
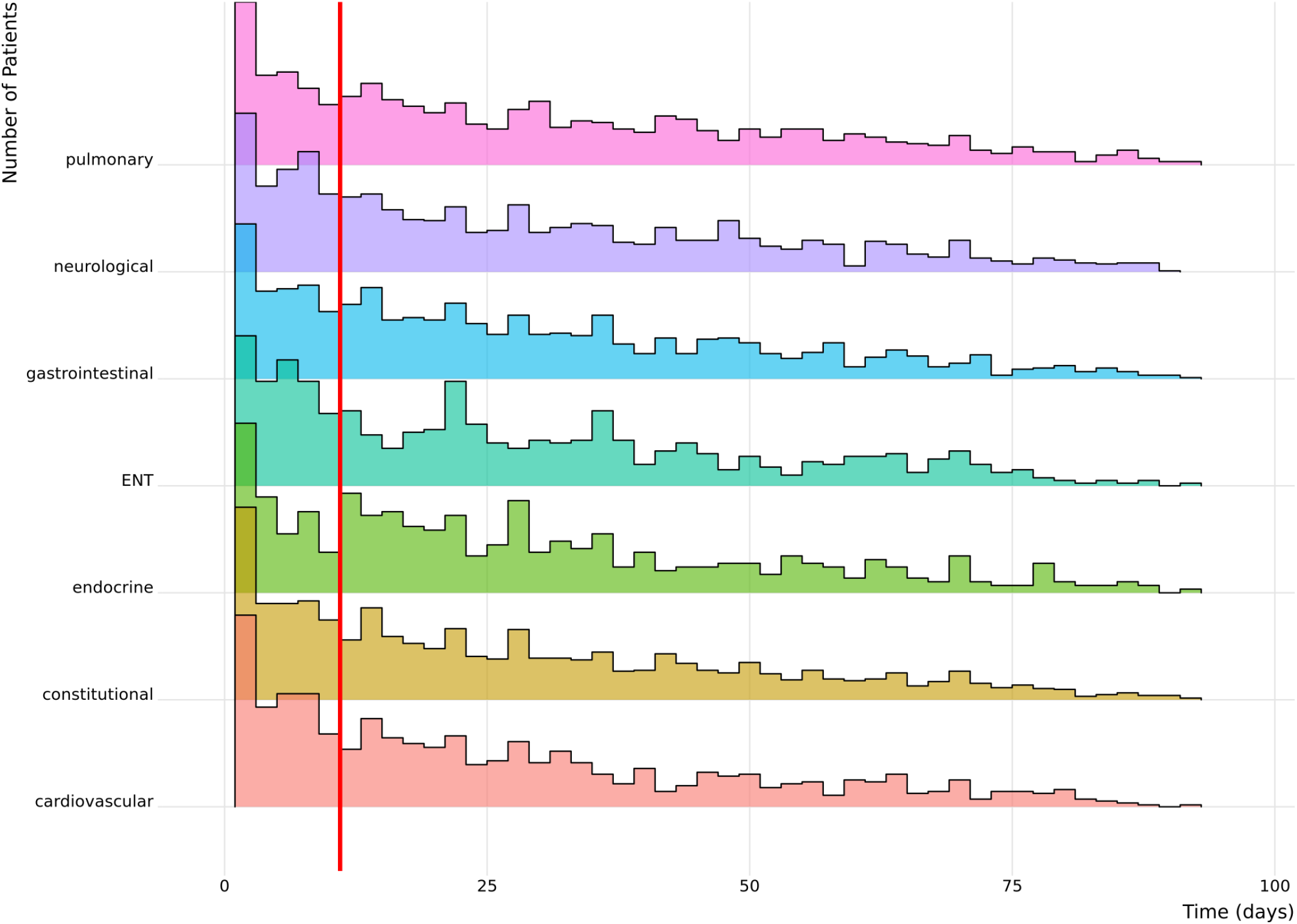
Time Between PASC-AM Diagnosis and Psychiatric Disease. Histogram showing the time between the first PASC-AM and mental disorder for each patient for each PASC-AM category. The X-axis shows the number of days between the first recorded PASC-AM for each patient and the recorded onset of psychiatric disease. A red line is drawn at eleven days to indicate data that was excluded in the sensitivity analysis where PASC-AMs occurring within 10 days before the first mental illness were not considered.

To understand the temporal relationship between PASC-AMs and new-onset psychiatric diseases, we selected patients with any PASCAM and a psychiatric outcome. We then found the time (in days) between each patient’s first PASC-AM in any category and diagnosis of psychiatric disease.

We then investigated the distribution of individual HPO terms for the seven categories described above. We compared counts of the observed HPO terms for each category among patients with diagnosis of new-onset anxiety, mood disorder, and psychosis. Statistical significance was assessed with a chi-squared test and adjusted for multiple testing. Only patients with at least one HPO term in the category were included in this analysis. To ensure that no patient was in multiple groups, we eliminated the category of all psychiatric diseases and removed patients who simultaneously presented with multiple outcomes, leaving 1,599,516 patients (Figure 5, Supplemental Figure S1 and Table S18).

**Figure 5:**
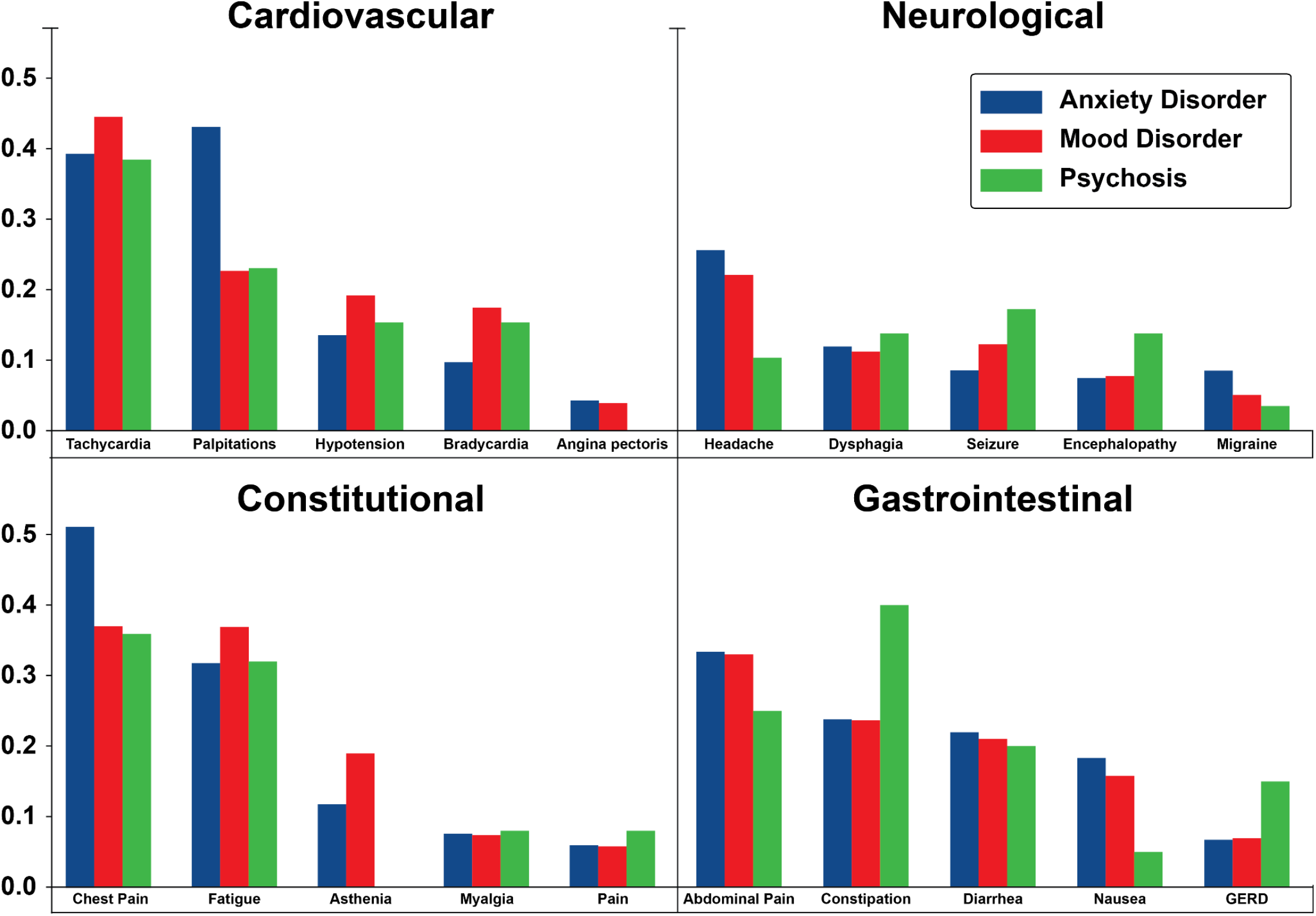
Proportion of Patients with HPO Feature by Category and Outcome. Here we examine the breakdown of individual HPO phenotypic features from symptom categories. The X-axis shows the five most prevalent HPO features from the displayed category. The Y-axis shows the proportion of patients with that feature from the set patients with the indicated HPO feature category and outcome. Significance of each finding was tested using chi-squared test (Table S18).

## Discussion

In this retrospective observational study on a cohort of 1,603,767 individuals following acute COVID-19 infection, we find that constitutional, neurological, gastrointestinal, cardiovascular and ENT PASC-AMs are associated with an increased incidence of new-onset psychiatric disease.

Five studies, including our own previous work, have performed large-scale EHR analyses that show significant and consistent but modest associations between SARS-CoV-2 infection and increased rates of psychiatric disorders.^5,6,9,22,23^ Differences in cumulative risk between patients who had COVID-19 and those who had other respiratory tract infections persist for at least 12 months. However, the risk of new-onset psychiatric disease decreases sharply after the first month after acute SARS-CoV-2 infection and two studies suggest the risk returns to baseline after 120 days.^6,9^ In contrast, a recent study showed that neuropsychiatric sequelae of severe COVID-19 infection were similar to those observed in other severe acute respiratory infections.^4^ With the exception of altered mental status, one study showed that neuropsychiatric manifestations are similar between patients with influenza and those with SARS-CoV2.^24^

Endocrine PASC-AMs were associated with a lower frequency of new-onset anxiety and mood disorders. It is not possible to make conclusions about causality in this retrospective study, but we speculate that behavioral manifestations observed in patients with endocrine disease may not lead to a diagnosis of primary psychiatric disease because of DSM-V guidelines.^25^ Since our study was restricted to diagnosis of new-onset primary psychiatric disease, this speculatively could explain the lower incidence.

Our results show that the presence of PASC-AMs in five clinical categories are associated with an increased incidence of newly diagnosed psychiatric disease following COVID-19. There are several possible explanations, including that pathophysiological mechanisms increase the risk of both new-onset psychiatric disease and PASC-AMs or that PASC-AMs themselves increase risk of newly diagnosed psychiatric disease following COVID-19. Our results show different PASC-AM category associations for anxiety, mood and psychotic disorders and that individual PASC-AMs within the categories contribute to these associations for each outcome. This may suggest there are several pathophysiological mechanisms^12^ in PASC that could explain the heterogeneity of phenotypic presentation.

### Study limitations

Our dataset is derived from over 65 institutions across the country with 5,858,748 cases of COVID-19, and thus is a representative sample of the COVID-19 positive population in the United States, but inconsistent or incomplete data collection could introduce biases. N3C employs a comprehensive suite of data quality checks to mitigate this problem,^26^ but residual issues cannot be ruled out. Retrospective analysis of EHR data does not allow any conclusions about pathomechanisms and cannot easily detect misdiagnoses.For example, individuals with postural orthostatic tachycardia syndrome (POTS) are sometimes mistakenly diagnosed with anxiety disorders such as panic disorder because of their tachycardia.^27^

### Conclusion

We have shown that the presence of PASC-AMs from any of five clinical categories is associated with increased incidence of newly diagnosed psychiatric disease. This is consistent with the association that has been described between clinical severity of several other chronic diseases (chronic heart failure, chronic kidney disease, chronic hepatitis C, and cancer) and psychiatric disease. In fact, that association is bidirectional: chronic disease is associated with increased rates of psychiatric disease, and psychiatric disease is associated with a higher risk of chronic disease occurrence, severity, or progression.^28–31^

Our results have important implications for both individual and public health. Timely and accurate diagnosis of psychiatric conditions has the potential to improve the quality of life for affected individuals; it may be worth systematically testing PASC patients for psychiatric sequelae. The scope of the COVID-19 pandemic is enormous, and it is essential to gain a deeper understanding of the natural history of PASC-related psychiatric diseases and their differential diagnoses to optimize care for affected individuals and institute appropriate public health measures.

## Data sharing

The data presented in this paper can be accessed upon application to the NCATS N3C Data Enclave at https://covid.cd2h.org/enclave.

**Acknowledgments**

The analyses described in this publication were conducted with data or tools accessed through the NCATS N3C Data Enclave (covid.cd2h.org/enclave) and supported by CD2H - The National COVID Cohort Collaborative (N3C) IDeA CTR Collaboration (3U24TR002306-04S2 NCATS U24 TR002306). This research was possible because of the patients whose information is included within the data from participating organizations (covid.cd2h.org/dtas) and the organizations and scientists (covid.cd2h.org/duas) who have contributed to the ongoing development of this community resource.^15^

We gratefully acknowledge the following core contributors to N3C:

Adam B. Wilcox, Adam M. Lee, Alexis Graves, Alfred (Jerrod) Anzalone, Amin Manna, Amit Saha, Amy Olex, Andrea Zhou, Andrew E. Williams, Andrew Southerland, Andrew T. Girvin, Anita Walden, Anjali A. Sharathkumar, Benjamin Amor, Benjamin Bates, Brian Hendricks, Brijesh Patel, Caleb Alexander, Carolyn Bramante, Cavin Ward-Caviness, Charisse Madlock-Brown, Christine Suver, Christopher Chute, Christopher Dillon, Chunlei Wu, Clare Schmitt, Cliff Takemoto, Dan Housman, Davera Gabriel, David A. Eichmann, Diego Mazzotti, Don Brown, Eilis Boudreau, Elaine Hill, Elizabeth Zampino, Emily Carlson Marti, Emily R. Pfaff, Evan French, Farrukh M Koraishy, Federico Mariona, Fred Prior, George Sokos, Greg Martin, Harold Lehmann, Heidi Spratt, Hemalkumar Mehta, Hongfang Liu, Hythem Sidky, J.W. Awori Hayanga, Jami Pincavitch, Jaylyn Clark, Jeremy Richard Harper, Jessica Islam, Jin Ge, Joel Gagnier, Joel H. Saltz, Joel Saltz, Johanna Loomba, John Buse, Jomol Mathew, Joni L. Rutter, Julie A. McMurry, Justin Guinney, Justin Starren, Karen Crowley, Katie Rebecca Bradwell, Kellie M. Walters, Ken Wilkins, Kenneth R. Gersing, Kenrick Dwain Cato, Kimberly Murray, Kristin Kostka, Lavance Northington, Lee Allan Pyles, Leonie Misquitta, Lesley Cottrell, Lili Portilla, Mariam Deacy, Mark M. Bissell, Marshall Clark, Mary Emmett, Mary Morrison Saltz, Matvey B. Palchuk, Melissa A. Haendel, Meredith Adams, Meredith Temple-O’Connor, Michael G. Kurilla, Michele Morris, Nabeel Qureshi, Nasia Safdar, Nicole Garbarini, Noha Sharafeldin, Ofer Sadan, Patricia A. Francis, Penny Wung Burgoon, Peter Robinson, Philip R.O. Payne, Rafael Fuentes, Randeep Jawa, Rebecca Erwin-Cohen, Rena Patel, Richard A. Moffitt, Richard L. Zhu, Rishi Kamaleswaran, Robert Hurley, Robert T. Miller, Saiju Pyarajan, Sam G. Michael, Samuel Bozzette, Sandeep Mallipattu, Satyanarayana Vedula, Scott Chapman, Shawn T. O’Neil, Soko Setoguchi, Stephanie S. Hong, Steve Johnson, Tellen D. Bennett, Tiffany Callahan, Umit Topaloglu, Usman Sheikh, Valery Gordon, Vignesh Subbian, Warren A. Kibbe, Wenndy Hernandez, Will Beasley, Will Cooper, William Hillegass, Xiaohan Tanner Zhang. Details of contributions available at covid.cd2h.org/core-contributors

## Disclaimer

**The content is solely the responsibility of the authors and does not necessarily represent the official views of the National Institutes of Health or the N3C program**.

### The following institutions whose data is released or pending

#### Available

Advocate Health Care Network — UL1TR002389: The Institute for Translational Medicine (ITM) • Boston University Medical Campus — UL1TR001430: Boston University Clinical and Translational Science Institute • Brown University — U54GM115677: Advance Clinical Translational Research (Advance-CTR) • Carilion Clinic — UL1TR003015: iTHRIV Integrated Translational health Research Institute of Virginia • Charleston Area Medical Center — U54GM104942: West Virginia Clinical and Translational Science Institute (WVCTSI) • Children’s Hospital Colorado — UL1TR002535: Colorado Clinical and Translational Sciences Institute • Columbia University Irving Medical Center — UL1TR001873: Irving Institute for Clinical and Translational Research • Duke University — UL1TR002553: Duke Clinical and Translational Science Institute • George Washington Children’s Research Institute — UL1TR001876: Clinical and Translational Science Institute at Children’s National (CTSA-CN) • George Washington University — UL1TR001876: Clinical and Translational Science Institute at Children’s National (CTSA-CN) • Indiana University School of Medicine — UL1TR002529: Indiana Clinical and Translational Science Institute • Johns Hopkins University — UL1TR003098: Johns Hopkins Institute for Clinical and Translational Research • Loyola Medicine — Loyola University Medical Center • Loyola University Medical Center — UL1TR002389: The Institute for Translational Medicine (ITM) • Maine Medical Center — U54GM115516: Northern New England Clinical & Translational Research (NNE-CTR) Network • Massachusetts General Brigham — UL1TR002541: Harvard Catalyst • Mayo Clinic Rochester — UL1TR002377: Mayo Clinic Center for Clinical and Translational Science (CCaTS) • Medical University of South Carolina — UL1TR001450: South Carolina Clinical & Translational Research Institute (SCTR) • Montefiore Medical Center — UL1TR002556: Institute for Clinical and Translational Research at Einstein and Montefiore • Nemours — U54GM104941: Delaware CTR ACCEL Program • NorthShore University HealthSystem — UL1TR002389: The Institute for Translational Medicine (ITM) • Northwestern University at Chicago — UL1TR001422: Northwestern University Clinical and Translational Science Institute (NUCATS) • OCHIN — INV-018455: Bill and Melinda Gates Foundation grant to Sage Bionetworks • Oregon Health & Science University — UL1TR002369: Oregon Clinical and Translational Research Institute • Penn State Health Milton S. Hershey Medical Center — UL1TR002014: Penn State Clinical and Translational Science Institute • Rush University Medical Center — UL1TR002389: The Institute for Translational Medicine (ITM) • Rutgers, The State University of New Jersey — UL1TR003017: New Jersey Alliance for Clinical and Translational Science • Stony Brook University — U24TR002306 • The Ohio State University — UL1TR002733: Center for Clinical and Translational Science • The State University of New York at Buffalo — UL1TR001412: Clinical and Translational Science Institute • The University of Chicago — UL1TR002389: The Institute for Translational Medicine (ITM) • The University of Iowa — UL1TR002537: Institute for Clinical and Translational Science • The University of Miami Leonard M. Miller School of Medicine — UL1TR002736: University of Miami Clinical and Translational Science Institute • The University of Michigan at Ann Arbor — UL1TR002240: Michigan Institute for Clinical and Health Research • The University of Texas Health Science Center at Houston — UL1TR003167: Center for Clinical and Translational Sciences (CCTS) • The University of Texas Medical Branch at Galveston — UL1TR001439: The Institute for Translational Sciences • The University of Utah — UL1TR002538: Uhealth Center for Clinical and Translational Science • Tufts Medical Center — UL1TR002544: Tufts Clinical and Translational Science Institute • Tulane University — UL1TR003096: Center for Clinical and Translational Science • University Medical Center New Orleans — U54GM104940: Louisiana Clinical and Translational Science (LA CaTS) Center • University of Alabama at Birmingham — UL1TR003096: Center for Clinical and Translational Science • University of Arkansas for Medical Sciences — UL1TR003107: UAMS Translational Research Institute • University of Cincinnati — UL1TR001425: Center for Clinical and Translational Science and Training • University of Colorado Denver, Anschutz Medical Campus — UL1TR002535: Colorado Clinical and Translational Sciences Institute • University of Illinois at Chicago — UL1TR002003: UIC Center for Clinical and Translational Science • University of Kansas Medical Center — UL1TR002366: Frontiers: University of Kansas Clinical and Translational Science Institute • University of Kentucky — UL1TR001998: UK Center for Clinical and Translational Science • University of Massachusetts Medical School Worcester — UL1TR001453: The UMass Center for Clinical and Translational Science (UMCCTS) • University of Minnesota — UL1TR002494: Clinical and Translational Science Institute • University of Mississippi Medical Center — U54GM115428: Mississippi Center for Clinical and Translational Research (CCTR) • University of Nebraska Medical Center — U54GM115458: Great Plains IDeA-Clinical & Translational Research • University of North Carolina at Chapel Hill — UL1TR002489: North Carolina Translational and Clinical Science Institute • University of Oklahoma Health Sciences Center — U54GM104938: Oklahoma Clinical and Translational Science Institute (OCTSI) • University of Rochester — UL1TR002001: UR Clinical & Translational Science Institute • University of Southern California — UL1TR001855: The Southern California Clinical and Translational Science Institute (SC CTSI) • University of Vermont — U54GM115516: Northern New England Clinical & Translational Research (NNE-CTR) Network • University of Virginia — UL1TR003015: iTHRIV Integrated Translational health Research Institute of Virginia • University of Washington — UL1TR002319: Institute of Translational Health Sciences • University of Wisconsin-Madison — UL1TR002373: UW Institute for Clinical and Translational Research • Vanderbilt University Medical Center — UL1TR002243: Vanderbilt Institute for Clinical and Translational Research • Virginia Commonwealth University — UL1TR002649: C. Kenneth and Dianne Wright Center for Clinical and Translational Research • Wake Forest University Health Sciences — UL1TR001420: Wake Forest Clinical and Translational Science Institute • Washington University in St. Louis — UL1TR002345: Institute of Clinical and Translational Sciences • Weill Medical College of Cornell University — UL1TR002384: Weill Cornell Medicine Clinical and Translational Science Center • West Virginia University — U54GM104942: West Virginia Clinical and Translational Science Institute (WVCTSI) Submitted: Icahn School of Medicine at Mount Sinai — UL1TR001433: ConduITS Institute for Translational Sciences • The University of Texas Health Science Center at Tyler — UL1TR003167: Center for Clinical and Translational Sciences (CCTS) • University of California, Davis — UL1TR001860: UCDavis Health Clinical and Translational Science Center • University of California, Irvine — UL1TR001414: The UC Irvine Institute for Clinical and Translational Science (ICTS) • University of California, Los Angeles — UL1TR001881: UCLA Clinical Translational Science Institute • University of California, San Diego — UL1TR001442: Altman Clinical and Translational Research Institute • University of California, San Francisco — UL1TR001872: UCSF Clinical and Translational Science Institute

#### Pending

Arkansas Children’s Hospital — UL1TR003107: UAMS Translational Research Institute • Baylor College of Medicine — None (Voluntary) • Children’s Hospital of Philadelphia — UL1TR001878: Institute for Translational Medicine and Therapeutics • Cincinnati Children’s Hospital Medical Center — UL1TR001425: Center for Clinical and Translational Science and Training • Emory University — UL1TR002378: Georgia Clinical and Translational Science Alliance • HonorHealth — None (Voluntary) • Loyola University Chicago — UL1TR002389: The Institute for Translational Medicine (ITM) • Medical College of Wisconsin — UL1TR001436: Clinical and Translational Science Institute of Southeast Wisconsin • MedStar Health Research Institute — UL1TR001409: The Georgetown-Howard Universities Center for Clinical and Translational Science (GHUCCTS) • MetroHealth — None (Voluntary) • Montana State University — U54GM115371: American Indian/Alaska Native CTR • NYU Langone Medical Center — UL1TR001445: Langone Health’s Clinical and Translational Science Institute • Ochsner Medical Center — U54GM104940: Louisiana Clinical and Translational Science (LA CaTS) Center • Regenstrief Institute — UL1TR002529: Indiana Clinical and Translational Science Institute • Sanford Research — None (Voluntary) • Stanford University — UL1TR003142: Spectrum: The Stanford Center for Clinical and Translational Research and Education • The Rockefeller University — UL1TR001866: Center for Clinical and Translational Science • The Scripps Research Institute — UL1TR002550: Scripps Research Translational Institute • University of Florida — UL1TR001427: UF Clinical and Translational Science Institute • University of New Mexico Health Sciences Center — UL1TR001449: University of New Mexico Clinical and Translational Science Center • University of Texas Health Science Center at San Antonio — UL1TR002645: Institute for Integration of Medicine and Science • Yale New Haven Hospital — UL1TR001863: Yale Center for Clinical Investigation

## Author contributions

Author Contributions: Dr Robinson and Dr Reese had full access to all of the data in the study and take responsibility for the integrity of the data and the accuracy of the data analysis.

Concept and design: Coleman, Casiraghi, Haendel, Reese, Robinson.

Acquisition, analysis, or interpretation of data: Coleman, Casiraghi, Callahan, Laraway, Haendel, Reese, Robinson

Drafting of the manuscript: Coleman, Casiraghi, Haendel, Reese, Robinson

Critical revision of the manuscript for important intellectual content: Coleman, Casiraghi, Blau, Chan, Davis, Clark, Seltzer, Valentini, Haendel, Reese, Robinson

Statistical analysis: Coleman, Casiraghi, Wilkins, Reese, Robinson Obtained funding: Haendel, Robinson.

Supervision: Reese, Robinson.

## References

1. Home - Johns Hopkins Coronavirus Resource Center [Internet]. [cited 2022 Jun 30]. Available from: https://coronavirus.jhu.edu/

2. Deer RR, Rock MA, Vasilevsky N, Carmody L, Rando H, Anzalone AJ, et al. Characterizing Long COVID: Deep Phenotype of a Complex Condition. EBioMedicine [Internet]. 2021 Dec;74:103722. Available from: http://dx.doi.org/10.1016/j.ebiom.2021.103722

3. Brodin P, Casari G, Townsend L, O’Farrelly C, Tancevski I, Löffler-Ragg J, et al. Studying severe long COVID to understand post-infectious disorders beyond COVID-19. Nat Med [Internet]. 2022 May;28(5):879–82. Available from: http://dx.doi.org/10.1038/s41591-022-01766-7

4. Clift AK, Ranger TA, Patone M, Coupland CAC, Hatch R, Thomas K, et al. Neuropsychiatric Ramifications of Severe COVID-19 and Other Severe Acute Respiratory Infections. JAMA Psychiatry [Internet]. 2022 May 11; Available from: http://dx.doi.org/10.1001/jamapsychiatry.2022.1067

5. Taquet M, Luciano S, Geddes JR, Harrison PJ. Bidirectional associations between COVID-19 and psychiatric disorder: retrospective cohort studies of 62 354 COVID-19 cases in the USA. Lancet Psychiatry [Internet]. 2020 Nov 9; Available from: http://dx.doi.org/10.1016/S2215-0366(20)30462-4

6. Coleman B, Casiraghi E, Blau H, Chan L, Haendel MA, Laraway B, et al. Risk of new-onset psychiatric sequelae of COVID-19 in the early and late post-acute phase. World Psychiatry [Internet]. 2022 Jun;21(2):319–20. Available from: http://dx.doi.org/10.1002/wps.20992

7. Castro VM, Rosand J, Giacino JT, McCoy TH, Perlis RH. Case-control study of neuropsychiatric symptoms in electronic health records following COVID-19 hospitalization in 2 academic health systems. Mol Psychiatry [Internet]. 2022 Jun 15; Available from: http://dx.doi.org/10.1038/s41380-022-01646-z

8. Taquet M, Geddes JR, Husain M, Luciano S, Harrison PJ. 6-month neurological and psychiatric outcomes in 236 379 survivors of COVID-19: a retrospective cohort study using electronic health records. Lancet Psychiatry [Internet]. 2021 May;8(5):416–27. Available from: http://dx.doi.org/10.1016/S2215-0366(21)00084-5

9. Murata F, Maeda M, Ishiguro C, Fukuda H. Acute and delayed psychiatric sequelae among patients hospitalised with COVID-19: a cohort study using LIFE study data. Gen Psych [Internet]. 2022 Jun 1 [cited 2022 Jun 29];35(3):e100802. Available from: https://gpsych.bmj.com/content/35/3/e100802.abstract

10. Spudich S, Nath A. Nervous system consequences of COVID-19. Science [Internet]. 2022 Jan 21;375(6578):267–9. Available from: http://dx.doi.org/10.1126/science.abm2052

11. Stefanou MI, Palaiodimou L, Bakola E, Smyrnis N, Papadopoulou M, Paraskevas GP, et al. Neurological manifestations of long-COVID syndrome: a narrative review. Ther Adv Chronic Dis [Internet]. 2022 Feb 17;13:20406223221076890. Available from: http://dx.doi.org/10.1177/20406223221076890

12. Mehandru S, Merad M. Pathological sequelae of long-haul COVID. Nat Immunol [Internet]. 2022 Feb;23(2):194–202. Available from: http://dx.doi.org/10.1038/s41590-021-01104-y

13. Reese, Blau, Bergquist, Loomba, Callahan, Laraway, et al. Generalizable Long COVID Subtypes: Findings from the NIH N3C and RECOVER Programs. medRxiv [Internet]. Available from: https://www.medrxiv.org/content/10.1101/2022.05.24.22275398v1.abstract

14. Robinson PN, Köhler S, Bauer S, Seelow D, Horn D, Mundlos S. The Human Phenotype Ontology: a tool for annotating and analyzing human hereditary disease. Am J Hum Genet. 2008;83(5):610–5.

15. Haendel MA, Chute CG, Bennett TD, Eichmann DA, Guinney J, Kibbe WA, et al. The National COVID Cohort Collaborative (N3C): Rationale, design, infrastructure, and deployment. J Am Med Inform Assoc [Internet]. 2021 Mar 1;28(3):427–43. Available from: http://dx.doi.org/10.1093/jamia/ocaa196

16. FitzHenry F, Resnic FS, Robbins SL, Denton J, Nookala L, Meeker D, et al. Creating a Common Data Model for Comparative Effectiveness with the Observational Medical Outcomes Partnership. Appl Clin Inform [Internet]. 2015 Aug 26;6(3):536–47. Available from: http://dx.doi.org/10.4338/ACI-2014-12-CR-0121

17. Köhler S, Gargano M, Matentzoglu N, Carmody LC, Lewis-Smith D, Vasilevsky NA, et al. The Human Phenotype Ontology in 2021. Nucleic Acids Res [Internet]. 2021 Jan 8;49(D1):D1207–17. Available from: http://dx.doi.org/10.1093/nar/gkaa1043

18. Callahan TJ, Stefanski AL, Wyrwa JM, Zeng C, Ostropolets A, Banda JM, et al. Ontologizing Health Systems Data at Scale: Making Translational Discovery a Reality [Internet]. arXiv [cs.DB]. 2022. Available from: http://arxiv.org/abs/2209.04732

19. Rubin DB. The Calculation of Posterior Distributions by Data Augmentation: Comment: A Noniterative Sampling/Importance Resampling Alternative to the Data Augmentation Algorithm for Creating a Few Imputations When Fractions of Missing Information Are Modest: The SIR Algorithm [Internet]. Vol. 82, Journal of the American Statistical Association. 1987. p. 543. Available from: http://dx.doi.org/10.2307/2289460

20. Casiraghi E, Wong R, Hall M, Coleman B, Notaro M, Evans MD, et al. A Methodological Framework for the Comparative Evaluation of Multiple Imputation Methods: Multiple Imputation of Race, Ethnicity and Body Mass Index in the U.S. National COVID Cohort Collaborative [Internet]. arXiv [cs.AI]. 2022. Available from: http://arxiv.org/abs/2206.06444

21. Gao M, Piernas C, Astbury NM, Hippisley-Cox J, O’Rahilly S, Aveyard P, et al. Associations between body-mass index and COVID-19 severity in 6·9 million people in England: a prospective, community-based, cohort study. Lancet Diabetes Endocrinol [Internet]. 2021 Jun;9(6):350–9. Available from: http://dx.doi.org/10.1016/S2213-8587(21)00089-9

22. Taquet M, Dercon Q, Luciano S, Geddes JR, Husain M, Harrison PJ. Incidence, co-occurrence, and evolution of long-COVID features: A 6-month retrospective cohort study of 273,618 survivors of COVID-19. PLoS Med [Internet]. 2021 Sep;18(9):e1003773. Available from: http://dx.doi.org/10.1371/journal.pmed.1003773

23. Xie Y, Xu E, Al-Aly Z. Risks of mental health outcomes in people with covid-19: cohort study. BMJ [Internet]. 2022 Feb 16;376:e068993. Available from: http://dx.doi.org/10.1136/bmj-2021-068993

24. Iosifescu AL, Hoogenboom WS, Buczek AJ, Fleysher R, Duong TQ. New‐onset and persistent neurological and psychiatric sequelae of COVID‐19 compared to influenza: A retrospective cohort study in a large New York City healthcare network [Internet]. International Journal of Methods in Psychiatric Research. 2022. Available from: http://dx.doi.org/10.1002/mpr.1914

25. American Psychiatric Association. Diagnostic and Statistical Manual of Mental Disorders (DSM-5®) [Internet]. American Psychiatric Pub; 2013. 991 p. Available from: https://play.google.com/store/books/details?id=-JivBAAAQBAJ

26. Pfaff ER, Girvin AT, Gabriel DL, Kostka K, Morris M, Palchuk M, et al. Synergies between Centralized and Federated Approaches to Data Quality: A Report from the National COVID Cohort Collaborative. J Am Med Inform Assoc [Internet]. 2021 Sep 30; Available from: http://dx.doi.org/10.1093/jamia/ocab217

27. Ståhlberg M, Reistam U, Fedorowski A, Villacorta H, Horiuchi Y, Bax J, et al. Post-COVID-19 Tachycardia Syndrome: A Distinct Phenotype of Post-Acute COVID-19 Syndrome. Am J Med [Internet]. 2021 Dec;134(12):1451–6. Available from: http://dx.doi.org/10.1016/j.amjmed.2021.07.004

28. Celano CM, Villegas AC, Albanese AM, Gaggin HK, Huffman JC. Depression and Anxiety in Heart Failure: A Review. Harv Rev Psychiatry [Internet]. 2018;26(4):175–84. Available from: http://dx.doi.org/10.1097/HRP.0000000000000162

29. Shirazian S, Grant CD, Aina O, Mattana J, Khorassani F, Ricardo AC. Depression in Chronic Kidney Disease and End-Stage Renal Disease: Similarities and Differences in Diagnosis, Epidemiology, and Management. Kidney Int Rep [Internet]. 2017 Jan;2(1):94–107. Available from: http://dx.doi.org/10.1016/j.ekir.2016.09.005

30. Huang X, Liu X, Yu Y. Depression and Chronic Liver Diseases: Are There Shared Underlying Mechanisms? Front Mol Neurosci [Internet]. 2017 May 8;10:134. Available from: http://dx.doi.org/10.3389/fnmol.2017.00134

31. Smith HR. Depression in cancer patients: Pathogenesis, implications and treatment (Review). Oncol Lett [Internet]. 2015 Apr;9(4):1509–14. Available from: http://dx.doi.org/10.3892/ol.2015.2944

