## Supplement for "Post-COVID Phenotypic Manifestations are Associated with New-Onset Psychiatric Disease: Findings from the NIH N3C and RECOVER Studies"

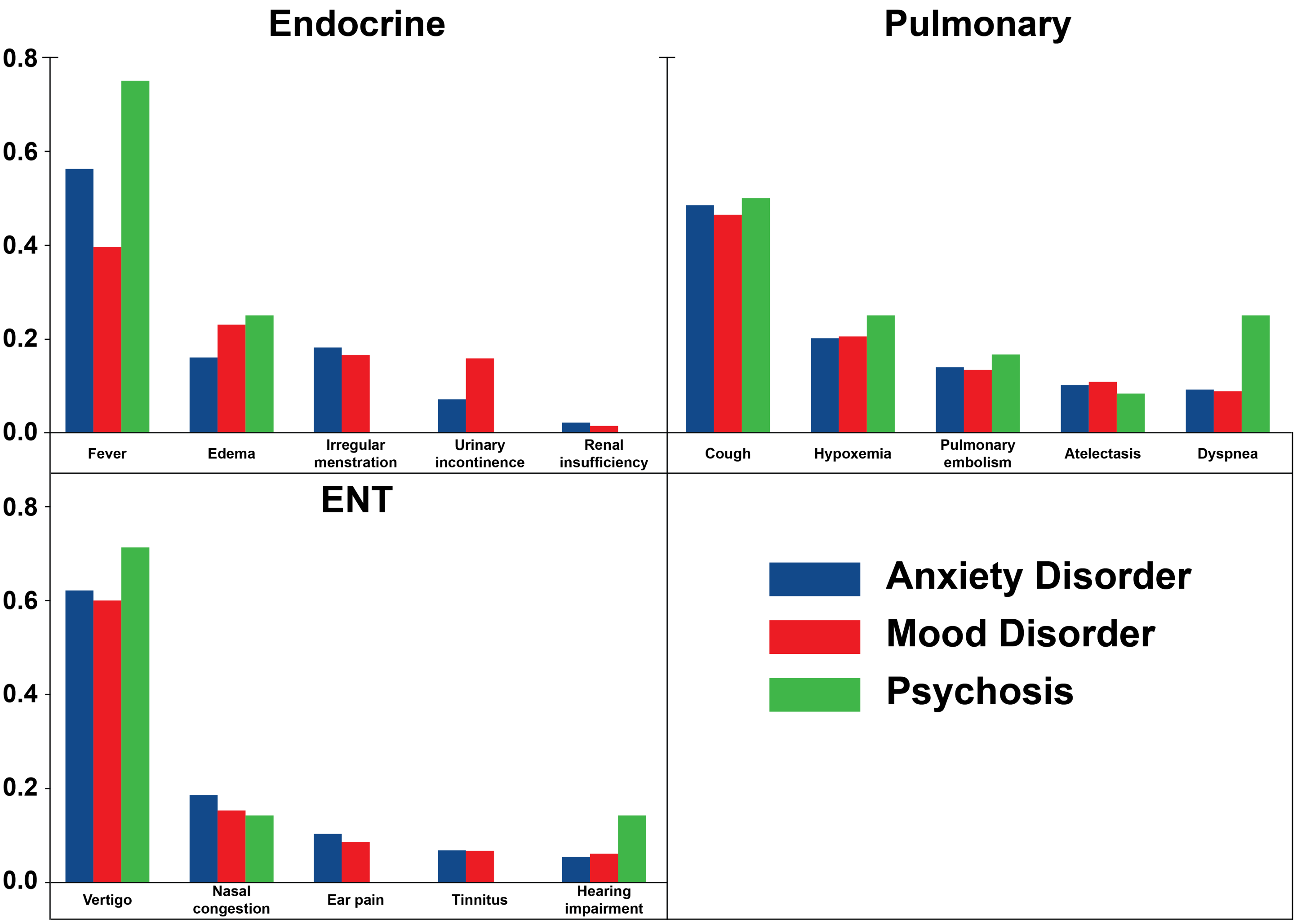


**Figure S1. Proportion of Patients with HPO Feature by Category and Outcome.** The X-axis shows the five most prevalent HPO features from the displayed category. The Y-axis shows the proportion of patients with that feature from the set patients with the indicated symptom category and outcome.

**Table S1. HPO Category: neuropsychiatric**. Given the removal of psychiatric HPO terms we refer to this category as **neurologic**. ✓: observed; ∉: no adequate mapping to OMOP; ×: excluded because it describes a psychiatric manifestation; ∅: not observed in the EHR dataset and therefore not used in the analysis.

| Id | label | status |
|---|---|---|
| HP:0033747 | Abnormal exteroceptive sensation | ∉ |
| HP:0031826 | Abnormal reflex | ✓ |
| HP:0100022 | Abnormality of movement | ✓ |
| HP:0041051 | Ageusia | ✓ |
| HP:0000718 | Aggressive behavior | ∅ |
| HP:0010524 | Agnosia | ✓ |
| HP:0030784 | Anomic aphasia | × |
| HP:0000458 | Anosmia | ✓ |
| HP:0033689 | Anterograde memory impairment | ∉ |
| HP:0000739 | Anxiety | ∅ |
| HP:0000741 | Apathy | ∅ |
| HP:0002381 | Aphasia | ✓ |
| HP:0001251 | Ataxia | ✓ |
| HP:0007018 | Attention deficit hyperactivity disorder | ∅ |
| HP:0008765 | Auditory hallucinations | ∅ |
| HP:0003487 | Babinski sign | ✓ |
| HP:0033849 | Bilingual aphasia | ∉ |
| HP:0002067 | Bradykinesia | ✓ |
| HP:0031843 | Bradyphrenia | × |
| HP:0033630 | Brain fog | ∉ |
| HP:0100543 | Cognitive impairment | ∉ |
| HP:0001289 | Confusion | ✓ |
| HP:0000746 | Delusions | ∅ |
| HP:0000716 | Depression | ∅ |
| HP:0031987 | Diminished ability to concentrate | ∅ |
| HP:0001260 | Dysarthria | ✓ |
| HP:0001310 | Dysmetria | ✓ |
| HP:0002015 | Dysphagia | ✓ |
| HP:0033838 | Dysphoria | ∉ |
| HP:0001332 | Dystonia | ✓ |
| HP:0000712 | Emotional lability | ∉ |
| HP:0001298 | Encephalopathy | ✓ |
| HP:0031844 | Euphoria | × |
| HP:0002427 | Expressive aphasia | ✓ |
| HP:0007209 | Facial paralysis | ∉ |
| HP:0000743 | Frontal release signs | ∉ |
| HP:0001288 | Gait disturbance | ∉ |
| HP:0000738 | Hallucinations | ∅ |
| HP:0030237 | Hand muscle weakness | ✓ |
| HP:0002315 | Headache | ✓ |
| HP:0100963 | Hyperesthesia | ✓ |
| HP:0002487 | Hyperkinetic movements | ✓ |
| HP:0033748 | Hypoesthesia | ∉ |

Continued on next page

**Table S1 – continued from previous page**

| Id | label | status |
|---|---|---|
| HP:0000224 | Hypogeusia | × |
| HP:0004409 | Hyposmia | ✓ |
| HP:0001252 | Hypotonia | ∉ |
| HP:0033051 | Impaired executive functioning | ✓ |
| HP:0100710 | Impulsivity | ∉ |
| HP:0100785 | Insomnia | ✓ |
| HP:0000737 | Irritability | ∅ |
| HP:0033688 | Long term memory impairment | ∉ |
| HP:0031355 | Maintenance insomnia | ∉ |
| HP:0100754 | Mania | ∅ |
| HP:0002354 | Memory impairment | ✓ |
| HP:0002076 | Migraine | ✓ |
| HP:0003394 | Muscle spasm | ✓ |
| HP:0001324 | Muscle weakness | ✓ |
| HP:0001278 | Orthostatic hypotension | ✓ |
| HP:0025269 | Panic attack | ∅ |
| HP:0031249 | Parageusia | ✓ |
| HP:0003401 | Paresthesia | ✓ |
| HP:0001300 | Parkinsonism | ✓ |
| HP:0033847 | Phantageusia | ∉ |
| HP:0033693 | Phantosmia | ∉ |
| HP:0002183 | Phonophobia | ∅ |
| HP:0001959 | Polydipsia | ∉ |
| HP:0001271 | Polyneuropathy | ✓ |
| HP:0033676 | Posttraumatic stress symptom | ∉ |
| HP:0033691 | Procedural memory loss | ∉ |
| HP:0033848 | Receptive aphasia | ∉ |
| HP:0012452 | Restless legs | ✓ |
| HP:0002063 | Rigidity | ∉ |
| HP:0001250 | Seizure | ✓ |
| HP:0033845 | Sense of impending doom | ∉ |
| HP:0000736 | Short attention span | ∅ |
| HP:0033687 | Short term memory impairment | ∉ |
| HP:0003202 | Skeletal muscle atrophy | ✓ |
| HP:0010535 | Sleep apnea | ✓ |
| HP:0002360 | Sleep disturbance | ✓ |
| HP:0031354 | Sleep onset insomnia | ∉ |
| HP:0001350 | Slurred speech | ✓ |
| HP:0003474 | Somatic sensory dysfunction | ✓ |
| HP:0001257 | Spasticity | ✓ |
| HP:0031589 | Suicidal ideation | ∉ |
| HP:0033844 | Tachyphrenia | ∉ |
| HP:0033694 | Tactile hallucination | ∉ |
| HP:0033705 | Tearfulness | ∉ |
| HP:0031356 | Terminal insomnia | × |
| HP:0001337 | Tremor | ✓ |
| HP:0012799 | Unilateral facial palsy | ∉ |

Continued on next page

**Table S1 – continued from previous page**

| Id | label | status |
|---|---|---|
| HP:0002367 | Visual hallucinations | ∅ |

**Table S2. HPO Category: Ear, Nose, and Throat (ENT).** ✓: observed; $\notin$: no adequate mapping to OMOP; $\times$: excluded because it describes a psychiatric manifestation; $\emptyset$: not observed in the EHR dataset and therefore not used in the analysis.

| Id | label | status |
|---|---|---|
| HP:0001618 | Dysphonia | $\notin$ |
| HP:0030766 | Ear pain | ✓ |
| HP:0000365 | Hearing impairment | ✓ |
| HP:0010780 | Hyperacusis | ✓ |
| HP:0001742 | Nasal congestion | ✓ |
| HP:0033050 | Pharyngalgia | ✓ |
| HP:0008629 | Pulsatile tinnitus | ✓ |
| HP:0012384 | Rhinitis | ✓ |
| HP:0000360 | Tinnitus | ✓ |
| HP:0002321 | Vertigo | ✓ |

**Table S3. HPO Category: skin**. ✓: observed; ∉: no adequate mapping to OMOP; ×: excluded because it describes a psychiatric manifestation; ∅: not observed in the EHR dataset and therefore not used in the analysis.

| **Id** | **label** | **status** |
|---|---|---|
| HP:0001596 | Alopecia | ✓ |
| HP:0011971 | Dermatographic urticaria | ✓ |
| HP:0000975 | Hyperhidrosis | ✓ |
| HP:0000967 | Petechiae | ✓ |
| HP:0000989 | Pruritus | ✓ |
| HP:0040189 | Scaling skin | ✓ |
| HP:0000988 | Skin rash | ✓ |

**Table S4. HPO Category: pulmonary**. ✓: observed; $\notin$: no adequate mapping to OMOP; ×: excluded because it describes a psychiatric manifestation; $\emptyset$: not observed in the EHR dataset and therefore not used in the analysis.

| Id | label | status |
|---|---|---|
| HP:0031983 | Abnormal pulmonary thoracic imaging finding | $\notin$ |
| HP:0006536 | Airway obstruction | $\notin$ |
| HP:0100750 | Atelectasis | ✓ |
| HP:0002110 | Bronchiectasis | ✓ |
| HP:0025180 | Centrilobular ground-glass opacification on pulmonary HRCT | $\notin$ |
| HP:0012735 | Cough | ✓ |
| HP:0033659 | Crazy-paving pattern | $\notin$ |
| HP:0045051 | Decreased DLCO | $\notin$ |
| HP:0033760 | Decreased maximal oxygen uptake | $\notin$ |
| HP:0033773 | Decreased RV/TLC ratio | $\notin$ |
| HP:0002094 | Dyspnea | ✓ |
| HP:0002875 | Exertional dyspnea | ✓ |
| HP:0025179 | Ground-glass opacification | $\notin$ |
| HP:0002105 | Hemoptysis | ✓ |
| HP:0012418 | Hypoxemia | ✓ |
| HP:0033709 | Increased sputum production | $\notin$ |
| HP:0030879 | Interlobular septal thickening | $\notin$ |
| HP:0031246 | Nonproductive cough | ✓ |
| HP:0030874 | Oxygen desaturation on exertion | $\notin$ |
| HP:0032177 | Parenchymal consolidation | $\notin$ |
| HP:0031944 | Pleural thickening | ✓ |
| HP:0033771 | Pleuritic chest pain | $\notin$ |
| HP:0002102 | Pleuritis | ✓ |
| HP:0031245 | Productive cough | ✓ |
| HP:0032446 | Pulmonary bulla | $\notin$ |
| HP:0002204 | Pulmonary embolism | ✓ |
| HP:0002206 | Pulmonary fibrosis | ✓ |
| HP:0033711 | Pulmonary interstitial thickening | $\notin$ |
| HP:0030877 | Reduced FEV1/FVC ratio | $\notin$ |
| HP:0032342 | Reduced forced expiratory volume in one second | $\notin$ |
| HP:0032341 | Reduced forced vital capacity | $\notin$ |
| HP:0033750 | Reduced functional residual capacity | $\notin$ |
| HP:0033753 | Reduced residual volume | $\notin$ |
| HP:0033169 | Reduced total lung capacity | $\notin$ |
| HP:0033710 | Rest dyspnea | $\notin$ |
| HP:0002091 | Restrictive ventilatory defect | ✓ |
| HP:0025390 | Reticular pattern on pulmonary HRCT | $\notin$ |
| HP:0031417 | Rhinorrhea | ✓ |
| HP:0030831 | Rhonchi | ✓ |
| HP:0025095 | Sneeze | ✓ |
| HP:0033609 | Solid pulmonary nodule | $\notin$ |
| HP:0033702 | Subpleural curvilinear line | $\notin$ |
| HP:0033610 | Subsolid pulmonary nodule | $\notin$ |
| HP:0002789 | Tachypnea | $\notin$ |

Continued on next page

**Table S4 – continued from previous page**

| Id | label | status |
|---|---|---|
| HP:0030828 | Wheezing | ✓ |

**Table S5. HPO Category: endocrine**. ✓: observed; ∉: no adequate mapping to OMOP; ×: excluded because it describes a psychiatric manifestation; ∅: not observed in the EHR dataset and therefore not used in the analysis.

| Id | label | status |
|---|---|---|
| HP:0012213 | Decreased glomerular filtration rate | ∉ |
| HP:0000819 | Diabetes mellitus | ✓ |
| HP:0000969 | Edema | ✓ |
| HP:0030014 | Female sexual dysfunction | ∉ |
| HP:0001945 | Fever | ✓ |
| HP:0002046 | Heat intolerance | ✓ |
| HP:0002045 | Hypothermia | ✓ |
| HP:0000858 | Irregular menstruation | ✓ |
| HP:0011134 | Low-grade fever | ✓ |
| HP:0040307 | Male sexual dysfunction | ✓ |
| HP:0000132 | Menorrhagia | ✓ |
| HP:0001733 | Pancreatitis | ✓ |
| HP:0033840 | Postmenopausal bleeding | ∉ |
| HP:0001954 | Recurrent fever | ✓ |
| HP:0000083 | Renal insufficiency | ✓ |
| HP:0005968 | Temperature instability | ∉ |
| HP:0033839 | Testicular pain | ∉ |
| HP:0000020 | Urinary incontinence | ✓ |

**Table S6. HPO Category: immunology**. ✓: observed; $\notin$: no adequate mapping to OMOP; ×: excluded because it describes a psychiatric manifestation; $\emptyset$: not observed in the EHR dataset and therefore not used in the analysis.

| Id | label | status |
|---|---|---|
| HP:0100845 | Anaphylactic shock | ✓ |
| HP:0032069 | Anti-thyroglobulin antibody positivity | $\notin$ |
| HP:0025379 | Anti-thyroid peroxidase antibody positivity | $\notin$ |
| HP:0003493 | Antinuclear antibody positivity | ✓ |
| HP:0002716 | Lymphadenopathy | ✓ |
| HP:0001888 | Lymphopenia | ✓ |

**Table S7. HPO Category: constitutional.** ✓: observed; ∉: no adequate mapping to OMOP; ×: excluded because it describes a psychiatric manifestation; ∅: not observed in the EHR dataset and therefore not used in the analysis.

| Id | label | status |
|---|---|---|
| HP:0002829 | Arthralgia | ✓ |
| HP:0001369 | Arthritis | ✓ |
| HP:0025406 | Asthenia | ✓ |
| HP:0033047 | Body ache | ∉ |
| HP:0002653 | Bone pain | ✓ |
| HP:0100749 | Chest pain | ✓ |
| HP:0031352 | Chest tightness | ✓ |
| HP:0025143 | Chills | ∉ |
| HP:0033850 | Coldness | ∉ |
| HP:0002355 | Difficulty walking | ✓ |
| HP:0033665 | Diminished health-related quality of life | ∉ |
| HP:0033667 | Diminished mental health | ∉ |
| HP:0033666 | Diminished physical functioning | ∉ |
| HP:0003546 | Exercise intolerance | ✓ |
| HP:0012378 | Fatigue | ✓ |
| HP:0033675 | Frailty | ∉ |
| HP:0031059 | Impaired ability to bathe oneself | ∉ |
| HP:0031060 | Impaired ability to dress oneself | ∉ |
| HP:0031058 | Impairment of activities of daily living | ∉ |
| HP:0033746 | Intrascapular pain | ∉ |
| HP:0009763 | Limb pain | ✓ |
| HP:0033834 | Malaise | ∉ |
| HP:0003326 | Myalgia | ✓ |
| HP:0033345 | Neuralgia | ∉ |
| HP:0030166 | Night sweats | ✓ |
| HP:0033695 | Occupational disability | ∉ |
| HP:0012531 | Pain | ✓ |
| HP:0030973 | Postexertional malaise | × |
| HP:0025144 | Shivering | ✓ |
| HP:0025258 | Stiff neck | ∉ |
| HP:0001824 | Weight loss | ∉ |
| HP:0000217 | Xerostomia | ✓ |

**Table S8. HPO Category: cardiovascular**. ✓: observed; ∉: no adequate mapping to OMOP; ×: excluded because it describes a psychiatric manifestation; ∅: not observed in the EHR dataset and therefore not used in the analysis.

| Id | label | status |
|---|---|---|
| HP:0001681 | Angina pectoris | ✓ |
| HP:0001662 | Bradycardia | ✓ |
| HP:4000006 | Elevated myocardial native T1 | ∉ |
| HP:4000003 | Elevated myocardial native T2 | ∉ |
| HP:0000822 | Hypertension | ✓ |
| HP:0002615 | Hypotension | ✓ |
| HP:0410173 | Increased circulating troponin I concentration | ∉ |
| HP:0410174 | Increased circulating troponin T concentration | ∉ |
| HP:0031862 | Increased heart rate variability | ∉ |
| HP:0033755 | Increased left ventricular end-diastolic volume | ∉ |
| HP:4000004 | Myocardial late gadolinium enhancement | ∉ |
| HP:0012819 | Myocarditis | ✓ |
| HP:0001962 | Palpitations | ✓ |
| HP:0001698 | Pericardial effusion | ✓ |
| HP:4000005 | Pericardial late gadolinium enhancement | ∉ |
| HP:0012664 | Reduced ejection fraction | ∉ |
| HP:0001297 | Stroke | ✓ |
| HP:0001279 | Syncope | ✓ |
| HP:0001649 | Tachycardia | ✓ |
| HP:0004936 | Venous thrombosis | ✓ |

**Table S9. HPO Category: gastrointestinal**. ✓: observed; $\notin$: no adequate mapping to OMOP; $\times$: excluded because it describes a psychiatric manifestation; $\emptyset$: not observed in the EHR dataset and therefore not used in the analysis.

| Id | label | status |
|---|---|---|
| HP:0002027 | Abdominal pain | ✓ |
| HP:0011458 | Abdominal symptom | $\notin$ |
| HP:0002039 | Anorexia | $\notin$ |
| HP:0002607 | Bowel incontinence | ✓ |
| HP:0002019 | Constipation | ✓ |
| HP:0002014 | Diarrhea | ✓ |
| HP:0033842 | Early satiety | $\notin$ |
| HP:0002592 | Gastric ulcer | ✓ |
| HP:0002020 | Gastroesophageal reflux | ✓ |
| HP:0002578 | Gastroparesis | ✓ |
| HP:0001397 | Hepatic steatosis | ✓ |
| HP:0012115 | Hepatitis | ✓ |
| HP:0002240 | Hepatomegaly | ✓ |
| HP:0004395 | Malnutrition | ✓ |
| HP:0002018 | Nausea | ✓ |
| HP:0033757 | Pancreatic steatosis | $\notin$ |
| HP:0004396 | Poor appetite | $\notin$ |
| HP:0001744 | Splenomegaly | ✓ |
| HP:0002013 | Vomiting | ✓ |

**Table S10. HPO Category: laboratory**. ✓: observed; ∉: no adequate mapping to OMOP; ×: excluded because it describes a psychiatric manifestation; ∅: not observed in the EHR dataset and therefore not used in the analysis.

| Id | label | status |
|---|---|---|
| HP:0012053 | Decreased circulating calcifediol concentration | ∉ |
| HP:0031964 | Elevated circulating alanine aminotransferase concentration | ∉ |
| HP:0003155 | Elevated circulating alkaline phosphatase concentration | ✓ |
| HP:0031956 | Elevated circulating aspartate aminotransferase concentration | ∉ |
| HP:0011227 | Elevated circulating C-reactive protein concentration | ✓ |
| HP:0003236 | Elevated circulating creatine kinase concentration | ∉ |
| HP:0003259 | Elevated circulating creatinine concentration | ✓ |
| HP:0033106 | Elevated circulating D-dimer concentration | ∉ |
| HP:0033833 | Elevated circulating soluble CD25 concentration | ∉ |
| HP:0002925 | Elevated circulating thyroid-stimulating hormone concentration | ✓ |
| HP:0003565 | Elevated erythrocyte sedimentation rate | ✓ |
| HP:0030948 | Elevated gamma-glutamyltransferase level | ∉ |
| HP:0003074 | Hyperglycemia | ✓ |
| HP:0002901 | Hypocalcemia | ✓ |
| HP:0011900 | Hypofibrinogenemia | × |
| HP:0001943 | Hypoglycemia | ✓ |
| HP:0002148 | Hypophosphatemia | ✓ |
| HP:0003281 | Increased circulating ferritin concentration | ∉ |
| HP:0030783 | Increased circulating interleukin 6 | ∉ |
| HP:0025435 | Increased circulating lactate dehydrogenase concentration | ∉ |
| HP:0031185 | Increased circulating NT-proBNP concentration | ∉ |
| HP:0032308 | Increased circulating procalcitonin concentration | ∉ |
| HP:0001873 | Thrombocytopenia | ✓ |

**Table S11. HPO Category: eye**. ✓: observed; $\notin$: no adequate mapping to OMOP; ×: excluded because it describes a psychiatric manifestation; $\emptyset$: not observed in the EHR dataset and therefore not used in the analysis.

| Id | label | status |
|---|---|---|
| HP:0000618 | Blindness | ✓ |
| HP:0000622 | Blurred vision | ✓ |
| HP:0000509 | Conjunctivitis | ✓ |
| HP:0000651 | Diplopia | ✓ |
| HP:0000640 | Gaze-evoked nystagmus | × |
| HP:0001097 | Keratoconjunctivitis sicca | ✓ |
| HP:0200026 | Ocular pain | ✓ |
| HP:0033841 | Ocular pruritus | $\notin$ |
| HP:0007994 | Peripheral visual field loss | × |
| HP:0000613 | Photophobia | ✓ |
| HP:0025337 | Red eye | ✓ |
| HP:0000572 | Visual loss | × |
| HP:0100832 | Vitreous floaters | ✓ |

**Table S12. Logistic Regression for All Factors and Outcomes.** Odds ratios, 95% confidence intervals, and p-values for the association of features in all factors and outcomes.

| | Any Mental Illness | | Anxiety Disorder | | Mood Disorder | | Psychosis | |
|---|---|---|---|---|---|---|---|---|
| Predictor | OR (CI) | p-val | OR (CI) | p-val | OR (CI) | p-val | OR (CI) | p-val |
| (Intercept) | 0.02 (0.02-0.03) | <0.05 | 0.02 (0.02-0.02) | <0.05 | 0.01 (0.01-0.01) | <0.05 | 0 (0-0) | <0.05 |
| constitutional | 1.26 (1.22-1.31) | <0.05 | 1.32 (1.26-1.38) | <0.05 | 1.17 (1.1-1.24) | <0.05 | 1.17 (0.87-1.58) | 0.31 |
| neurological | 1.21 (1.16-1.25) | <0.05 | 1.16 (1.11-1.22) | <0.05 | 1.27 (1.2-1.34) | <0.05 | 1.4 (1.1-1.78) | <0.05 |
| gastrointestinal | 1.13 (1.09-1.17) | <0.05 | 1.16 (1.11-1.22) | <0.05 | 1.09 (1.03-1.16) | <0.05 | 1.16 (0.86-1.57) | 0.34 |
| pulmonary | 1.05 (1-1.1) | 0.057 | 1.03 (0.97-1.09) | 0.4 | 1.07 (0.99-1.16) | 0.071 | 0.96 (0.64-1.43) | 0.84 |
| cardiovascular | 1.35 (1.27-1.42) | <0.05 | 1.45 (1.35-1.54) | <0.05 | 1.07 (0.97-1.18) | 0.19 | 1.6 (1.1-2.35) | <0.05 |
| ENT | 1.07 (1-1.15) | 0.05 | 1.16 (1.07-1.26) | <0.05 | 0.94 (0.83-1.05) | 0.28 | 0.87 (0.45-1.65) | 0.66 |
| endocrine | 0.84 (0.77-0.92) | <0.05 | 0.86 (0.77-0.95) | <0.05 | 0.87 (0.76-0.99) | <0.05 | 0.4 (0.15-1.07) | 0.067 |
| BMI under 20 | 0.57 (0.53-0.63) | <0.05 | 0.62 (0.57-0.68) | <0.05 | 0.44 (0.39-0.51) | <0.05 | 0.38 (0.19-0.79) | <0.05 |
| BMI 20 to 25 | 0.85 (0.79-0.91) | <0.05 | 0.87 (0.8-0.94) | <0.05 | 0.78 (0.72-0.86) | <0.05 | 1.03 (0.64-1.68) | 0.89 |
| BMI 25 to 30 | 0.85 (0.79-0.91) | <0.05 | 0.86 (0.8-0.93) | <0.05 | 0.8 (0.73-0.86) | <0.05 | 1 (0.64-1.55) | 0.99 |
| BMI 30 to 35 | 0.88 (0.82-0.95) | <0.05 | 0.89 (0.82-0.97) | <0.05 | 0.84 (0.76-0.92) | <0.05 | 1.13 (0.7-1.81) | 0.61 |
| BMI 35 to 40 | 0.91 (0.84-0.99) | <0.05 | 0.91 (0.83-0.99) | <0.05 | 0.89 (0.8-1) | <0.05 | 0.99 (0.59-1.68) | 0.98 |
| gender MALE | 0.54 (0.52-0.55) | <0.05 | 0.52 (0.5-0.54) | <0.05 | 0.51 (0.49-0.53) | <0.05 | 1.13 (0.92-1.37) | 0.24 |
| gender FEMALE | 1 | REF | 1 | REF | 1 | REF | 1 | REF |
| smoking status | 1.33 (1.22-1.45) | <0.05 | 1.31 (1.18-1.46) | <0.05 | 1.32 (1.16-1.5) | <0.05 | 1.62 (0.92-2.88) | 0.096 |
| race ethnicity White Non Hispanic | 1 | REF | 1 | REF | 1 | REF | 1 | REF |
| race ethnicity Other Non Hispanic | 0.66 (0.59-0.73) | <0.05 | 0.63 (0.55-0.72) | <0.05 | 0.66 (0.56-0.77) | <0.05 | 1.08 (0.48-2.42) | 0.86 |
| race ethnicity Native Hawaiian or Other Pacific Islander Non Hispanic | 0.67 (0.45-1) | 0.05 | 0.62 (0.38-1.02) | 0.059 | 0.71 (0.4-1.26) | 0.25 | null | null |
| race ethnicity Hispanic or Latino Any Race | 1.03 (0.99-1.07) | 0.15 | 1 (0.95-1.04) | 0.88 | 1 (0.95-1.06) | 0.88 | 1.25 (0.91-1.72) | 0.17 |
| race ethnicity Unknown | 0.86 (0.81-0.92) | <0.05 | 0.86 (0.8-0.93) | <0.05 | 0.82 (0.74-0.91) | <0.05 | 1.1 (0.65-1.86) | 0.72 |
| race ethnicity Black or African American Non Hispanic | 0.83 (0.8-0.86) | <0.05 | 0.75 (0.72-0.78) | <0.05 | 0.87 (0.82-0.91) | <0.05 | 2.33 (1.85-2.93) | <0.05 |
| race ethnicity Asian Non Hispanic | 0.69 (0.63-0.75) | <0.05 | 0.65 (0.58-0.72) | <0.05 | 0.65 (0.57-0.75) | <0.05 | 1.12 (0.57-2.19) | 0.74 |
| chronic resp | 1.29 (1.24-1.33) | <0.05 | 1.33 (1.27-1.38) | <0.05 | 1.19 (1.13-1.26) | <0.05 | 0.99 (0.73-1.35) | 0.95 |
| diabetes2 | 1 (0.96-1.05) | 0.88 | 0.92 (0.87-0.97) | <0.05 | 1.11 (1.05-1.19) | <0.05 | 0.98 (0.7-1.36) | 0.88 |
| hepatic steatosis | 1.09 (1-1.17) | <0.05 | 1.12 (1.02-1.24) | <0.05 | 1.08 (0.96-1.21) | 0.19 | 0.16 (0.04-0.65) | <0.05 |
| hypertension | 1.12 (1.08-1.16) | <0.05 | 1.06 (1.02-1.11) | <0.05 | 1.11 (1.06-1.17) | <0.05 | 0.99 (0.75-1.3) | 0.93 |
| hypertensive kidney disease | 1.03 (0.93-1.15) | 0.54 | 1.05 (0.92-1.21) | 0.47 | 1 (0.87-1.16) | 0.98 | 0.92 (0.46-1.85) | 0.82 |
| ischemic heart disease | 0.95 (0.88-1.02) | 0.17 | 0.9 (0.82-0.98) | <0.05 | 1.08 (0.97-1.19) | 0.16 | 0.85 (0.52-1.4) | 0.53 |
| neoplasm | 1.08 (1.05-1.12) | <0.05 | 1.09 (1.05-1.14) | <0.05 | 1 (0.95-1.05) | 0.94 | 0.73 (0.55-0.99) | <0.05 |
| nicotine dependence | 1.4 (1.31-1.51) | <0.05 | 1.41 (1.3-1.53) | <0.05 | 1.5 (1.36-1.66) | <0.05 | 1.75 (1.08-2.83) | <0.05 |
| non hypertensive chronic kidney disease | 0.99 (0.92-1.07) | 0.76 | 0.86 (0.78-0.95) | <0.05 | 1.15 (1.04-1.28) | <0.05 | 1.21 (0.72-2.01) | 0.47 |
| non ischemic heart disease | 1.03 (0.99-1.07) | 0.18 | 0.99 (0.94-1.04) | 0.6 | 1.04 (0.98-1.1) | 0.24 | 1.69 (1.26-2.26) | <0.05 |
| other liver disease | 1.2 (1.11-1.29) | <0.05 | 1.15 (1.05-1.26) | <0.05 | 1.23 (1.11-1.37) | <0.05 | 1.9 (1.17-3.09) | <0.05 |

**Table S13. Logistic Regression for All Factors and Outcomes for Outpatients Only.** To rule out our results being an artifact from mixing patients with inpatient and outpatient status, we repeated the analysis by separating inpatients and outpatients. Odds ratios, 95% confidence intervals, and p-values for the association of features in all factors and outcomes.

| | Any Mental Illness | | Anxiety Disorder | | Mood Disorder | | Psychosis | |
|---|---|---|---|---|---|---|---|---|
| Predictor | OR (CI) | p-val | OR (CI) | p-val | OR (CI) | p-val | OR (CI) | p-val |
| (Intercept) | 0.02 (0.02-0.03) | <0.05 | 0.02 (0.02-0.02) | <0.05 | 0.01 (0.01-0.01) | <0.05 | 0 (0-0) | <0.05 |
| constitutional | 1.29 (1.24-1.35) | <0.05 | 1.34 (1.28-1.41) | <0.05 | 1.19 (1.12-1.27) | <0.05 | 1.42 (1.03-1.95) | <0.05 |
| neurological | 1.2 (1.15-1.25) | <0.05 | 1.17 (1.11-1.23) | <0.05 | 1.26 (1.18-1.33) | <0.05 | 1.19 (0.86-1.65) | 0.3 |
| gastrointestinal | 1.13 (1.08-1.17) | <0.05 | 1.16 (1.11-1.22) | <0.05 | 1.09 (1.02-1.16) | <0.05 | 1.24 (0.89-1.72) | 0.2 |
| pulmonary | 1.01 (0.95-1.07) | 0.71 | 0.99 (0.92-1.07) | 0.83 | 1.04 (0.95-1.14) | 0.42 | 0.94 (0.57-1.57) | 0.82 |
| cardiovascular | 1.34 (1.26-1.43) | <0.05 | 1.47 (1.36-1.58) | <0.05 | 1.02 (0.91-1.14) | 0.79 | 1.62 (1.03-2.56) | <0.05 |
| ENT | 1.1 (1.02-1.19) | <0.05 | 1.2 (1.1-1.3) | <0.05 | 0.95 (0.84-1.07) | 0.4 | 0.85 (0.41-1.77) | 0.67 |
| endocrine | 0.81 (0.73-0.9) | <0.05 | 0.83 (0.74-0.93) | <0.05 | 0.84 (0.72-0.98) | <0.05 | 0.29 (0.07-1.15) | 0.079 |
| BMI under 20 | 0.58 (0.53-0.63) | <0.05 | 0.63 (0.57-0.69) | <0.05 | 0.45 (0.39-0.51) | <0.05 | 0.38 (0.18-0.8) | <0.05 |
| BMI 20 to 25 | 0.86 (0.79-0.93) | <0.05 | 0.88 (0.8-0.96) | <0.05 | 0.79 (0.72-0.87) | <0.05 | 1.03 (0.61-1.72) | 0.92 |
| BMI 25 to 30 | 0.86 (0.79-0.92) | <0.05 | 0.87 (0.8-0.95) | <0.05 | 0.8 (0.73-0.87) | <0.05 | 1 (0.62-1.6) | 0.99 |
| BMI 30 to 35 | 0.89 (0.82-0.96) | <0.05 | 0.9 (0.83-0.98) | <0.05 | 0.84 (0.76-0.93) | <0.05 | 1.09 (0.66-1.8) | 0.73 |
| BMI 35 to 40 | 0.92 (0.84-1) | <0.05 | 0.91 (0.83-1) | 0.06 | 0.9 (0.81-1.01) | 0.079 | 0.93 (0.53-1.61) | 0.78 |
| gender MALE | 0.52 (0.51-0.54) | <0.05 | 0.5 (0.49-0.52) | <0.05 | 0.49 (0.47-0.52) | <0.05 | 1.26 (1.02-1.56) | <0.05 |
| gender FEMALE | 1 | REF | 1 | REF | 1 | REF | 1 | REF |
| smoking status | 1.33 (1.21-1.46) | <0.05 | 1.3 (1.16-1.46) | <0.05 | 1.33 (1.16-1.52) | <0.05 | 1.7 (0.91-3.16) | 0.097 |
| race ethnicity White Non Hispanic | 1 | REF | 1 | REF | 1 | REF | 1 | REF |
| race ethnicity Other Non Hispanic | 0.66 (0.58-0.73) | <0.05 | 0.62 (0.54-0.71) | <0.05 | 0.66 (0.56-0.78) | <0.05 | 1.17 (0.52-2.64) | 0.7 |
| race ethnicity Native Hawaiian or Other Pacific Islander Non Hispanic | 0.7 (0.47-1.06) | 0.093 | 0.66 (0.4-1.1) | 0.11 | 0.74 (0.41-1.34) | 0.33 | null | null |
| race ethnicity Hispanic or Latino Any Race | 1.03 (0.99-1.07) | 0.12 | 1 (0.96-1.05) | 0.99 | 1.01 (0.96-1.07) | 0.63 | 1.27 (0.91-1.77) | 0.17 |
| race ethnicity Unknown | 0.86 (0.81-0.93) | <0.05 | 0.88 (0.81-0.95) | <0.05 | 0.82 (0.74-0.91) | <0.05 | 0.94 (0.51-1.73) | 0.85 |
| race ethnicity Black or African American Non Hispanic | 0.82 (0.79-0.85) | <0.05 | 0.73 (0.7-0.77) | <0.05 | 0.87 (0.82-0.92) | <0.05 | 2.38 (1.86-3.04) | <0.05 |
| race ethnicity Asian Non Hispanic | 0.68 (0.62-0.75) | <0.05 | 0.64 (0.58-0.72) | <0.05 | 0.64 (0.55-0.74) | <0.05 | 1.13 (0.56-2.3) | 0.74 |
| chronic resp | 1.31 (1.26-1.36) | <0.05 | 1.34 (1.29-1.41) | <0.05 | 1.21 (1.14-1.29) | <0.05 | 0.94 (0.66-1.32) | 0.71 |
| diabetes2 | 0.97 (0.93-1.02) | 0.23 | 0.87 (0.82-0.93) | <0.05 | 1.09 (1.02-1.17) | <0.05 | 1.09 (0.75-1.57) | 0.65 |
| hepatic steatosis | 1.08 (1-1.18) | 0.064 | 1.12 (1.01-1.24) | <0.05 | 1.08 (0.95-1.22) | 0.24 | 0.09 (0.01-0.67) | <0.05 |
| hypertension | 1.11 (1.07-1.15) | <0.05 | 1.07 (1.02-1.11) | <0.05 | 1.11 (1.05-1.17) | <0.05 | 0.96 (0.71-1.3) | 0.8 |
| hypertensive kidney disease | 1.09 (0.97-1.24) | 0.15 | 1.11 (0.94-1.31) | 0.22 | 1.03 (0.87-1.22) | 0.73 | 0.73 (0.28-1.87) | 0.51 |
| ischemic heart disease | 0.98 (0.9-1.06) | 0.58 | 0.93 (0.83-1.03) | 0.16 | 1.09 (0.97-1.22) | 0.17 | 0.81 (0.44-1.48) | 0.49 |
| neoplasm | 1.08 (1.05-1.12) | <0.05 | 1.09 (1.04-1.13) | <0.05 | 1 (0.95-1.06) | 0.97 | 0.65 (0.46-0.91) | <0.05 |
| nicotine dependence | 1.42 (1.32-1.53) | <0.05 | 1.44 (1.31-1.57) | <0.05 | 1.51 (1.36-1.68) | <0.05 | 1.82 (1.08-3.06) | <0.05 |
| non hypertensive chronic kidney disease | 0.97 (0.89-1.06) | 0.56 | 0.81 (0.72-0.91) | <0.05 | 1.22 (1.08-1.37) | <0.05 | 1.08 (0.58-2.01) | 0.81 |
| non ischemic heart disease | 1.01 (0.97-1.06) | 0.6 | 0.97 (0.92-1.03) | 0.31 | 1.03 (0.96-1.1) | 0.4 | 1.64 (1.18-2.27) | <0.05 |
| other liver disease | 1.2 (1.11-1.3) | <0.05 | 1.17 (1.06-1.29) | <0.05 | 1.22 (1.09-1.38) | <0.05 | 2.31 (1.37-3.91) | <0.05 |

**Table S14. Logistic Regression for All Factors and Outcomes for Inpatients Only.** To rule out our results being an artifact from mixing patients with inpatient and outpatient status, we repeated the analysis by separating inpatients and outpatients. Odds ratios, 95% confidence intervals, and p-values for the association of features in all factors and outcomes.

| | Any Mental Illness | | Anxiety Disorder | | Mood Disorder | | Psychosis | |
|---|---|---|---|---|---|---|---|---|
| Predictor | OR (CI) | p-val | OR (CI) | p-val | OR (CI) | p-val | OR (CI) | p-val |
| (Intercept) | 0.02 (0.02-0.03) | <0.05 | 0.02 (0.01-0.02) | <0.05 | 0.01 (0.01-0.01) | <0.05 | 0 (0-0) | <0.05 |
| constitutional | 1.11 (1.01-1.22) | <0.05 | 1.19 (1.07-1.33) | <0.05 | 1.07 (0.93-1.23) | 0.34 | 0.5 (0.21-1.18) | 0.11 |
| neurological | 1.19 (1.1-1.28) | <0.05 | 1.11 (1-1.23) | <0.05 | 1.26 (1.13-1.4) | <0.05 | 1.85 (1.3-2.63) | <0.05 |
| gastrointestinal | 1.13 (1.03-1.25) | <0.05 | 1.17 (1.05-1.32) | <0.05 | 1.09 (0.94-1.26) | 0.25 | 0.89 (0.43-1.83) | 0.75 |
| pulmonary | 1.08 (0.99-1.19) | 0.092 | 1.07 (0.95-1.2) | 0.27 | 1.08 (0.94-1.24) | 0.28 | 0.88 (0.45-1.73) | 0.72 |
| cardiovascular | 1.31 (1.16-1.47) | <0.05 | 1.31 (1.13-1.52) | <0.05 | 1.17 (0.97-1.41) | 0.096 | 1.5 (0.74-3.03) | 0.27 |
| ENT | 0.91 (0.72-1.14) | 0.4 | 0.94 (0.71-1.24) | 0.65 | 0.91 (0.65-1.28) | 0.6 | 0.93 (0.24-3.66) | 0.92 |
| endocrine | 0.91 (0.75-1.11) | 0.34 | 0.95 (0.75-1.2) | 0.66 | 0.94 (0.71-1.25) | 0.68 | 0.67 (0.16-2.8) | 0.59 |
| BMI under 20 | 0.8 (0.59-1.1) | 0.17 | 0.89 (0.62-1.27) | 0.51 | 0.66 (0.4-1.11) | 0.12 | 0.78 (0.09-6.96) | 0.83 |
| BMI 20 to 25 | 0.8 (0.67-0.95) | <0.05 | 0.78 (0.62-0.98) | <0.05 | 0.79 (0.6-1.04) | 0.091 | 1.34 (0.39-4.6) | 0.64 |
| BMI 25 to 30 | 0.83 (0.7-0.98) | <0.05 | 0.8 (0.64-0.99) | <0.05 | 0.83 (0.65-1.06) | 0.14 | 1.09 (0.35-3.38) | 0.88 |
| BMI 30 to 35 | 0.86 (0.74-0.99) | <0.05 | 0.85 (0.7-1.02) | 0.073 | 0.82 (0.67-1) | <0.05 | 1.53 (0.55-4.26) | 0.41 |
| BMI 35 to 40 | 0.86 (0.71-1.05) | 0.13 | 0.86 (0.68-1.09) | 0.22 | 0.82 (0.62-1.08) | 0.16 | 1.5 (0.46-4.91) | 0.5 |
| gender MALE | 1 | REF | 1 | REF | 1 | REF | 1 | REF |
| gender FEMALE | 1.38 (1.26-1.51) | <0.05 | 1.36 (1.22-1.52) | <0.05 | 1.48 (1.3-1.69) | <0.05 | 2.29 (1.28-4.11) | <0.05 |
| smoking status | 1.26 (0.99-1.61) | 0.055 | 1.35 (1-1.82) | 0.05 | 1.19 (0.84-1.69) | 0.32 | 1.35 (0.33-5.57) | 0.68 |
| race ethnicity White Non Hispanic | 1 | REF | 1 | REF | 1 | REF | 1 | REF |
| race ethnicity Other Non Hispanic | 0.75 (0.45-1.23) | 0.25 | 0.93 (0.54-1.62) | 0.81 | 0.6 (0.27-1.34) | 0.21 | null | null |
| race ethnicity Native Hawaiian or Other Pacific Islander Non Hispanic | 0.44 (0.11-1.77) | 0.25 | 0.34 (0.05-2.44) | 0.28 | 0.46 (0.06-3.32) | 0.44 | null | null |
| race ethnicity Hispanic or Latino Any Race | 0.91 (0.79-1.04) | 0.16 | 0.89 (0.75-1.05) | 0.18 | 0.82 (0.67-1) | 0.055 | 0.94 (0.35-2.48) | 0.89 |
| race ethnicity Unknown | 0.71 (0.58-0.87) | <0.05 | 0.65 (0.5-0.84) | <0.05 | 0.71 (0.52-0.96) | <0.05 | 1.47 (0.51-4.28) | 0.48 |
| race ethnicity Black or African American Non Hispanic | 0.84 (0.75-0.93) | <0.05 | 0.83 (0.72-0.95) | <0.05 | 0.78 (0.66-0.92) | <0.05 | 1.69 (0.9-3.17) | 0.1 |
| race ethnicity Asian Non Hispanic | 0.68 (0.5-0.93) | <0.05 | 0.66 (0.44-0.98) | <0.05 | 0.7 (0.45-1.1) | 0.12 | 0.87 (0.12-6.57) | 0.9 |
| chronic resp | 1.1 (0.99-1.24) | 0.088 | 1.14 (0.99-1.32) | 0.063 | 1.05 (0.89-1.25) | 0.57 | 1.2 (0.61-2.38) | 0.6 |
| diabetes2 | 1.1 (0.99-1.22) | 0.08 | 1.08 (0.94-1.23) | 0.27 | 1.18 (1.01-1.37) | <0.05 | 0.6 (0.29-1.22) | 0.16 |
| hepatic steatosis | 1.13 (0.91-1.39) | 0.27 | 1.18 (0.91-1.54) | 0.21 | 1.13 (0.82-1.54) | 0.46 | 0.53 (0.07-4.05) | 0.54 |
| hypertension | 1.05 (0.95-1.16) | 0.37 | 0.99 (0.87-1.12) | 0.83 | 1.06 (0.91-1.23) | 0.48 | 0.88 (0.46-1.7) | 0.71 |
| hypertensive kidney disease | 0.91 (0.75-1.12) | 0.37 | 0.92 (0.71-1.18) | 0.5 | 0.99 (0.74-1.34) | 0.97 | 1.31 (0.45-3.76) | 0.62 |
| ischemic heart disease | 0.85 (0.73-0.99) | <0.05 | 0.77 (0.63-0.94) | <0.05 | 1.04 (0.84-1.3) | 0.7 | 1 (0.42-2.42) | 0.99 |
| neoplasm | 1.08 (0.97-1.2) | 0.15 | 1.12 (0.98-1.27) | 0.088 | 0.96 (0.82-1.12) | 0.61 | 1.28 (0.68-2.39) | 0.44 |
| nicotine dependence | 1.3 (1.06-1.6) | <0.05 | 1.23 (0.95-1.59) | 0.12 | 1.42 (1.06-1.91) | <0.05 | 1.39 (0.4-4.79) | 0.6 |
| non hypertensive chronic kidney disease | 0.92 (0.79-1.07) | 0.28 | 0.93 (0.76-1.13) | 0.45 | 0.85 (0.67-1.07) | 0.17 | 1.36 (0.55-3.35) | 0.5 |
| non ischemic heart disease | 1.01 (0.9-1.12) | 0.91 | 0.97 (0.85-1.11) | 0.64 | 0.98 (0.83-1.15) | 0.76 | 1.47 (0.75-2.88) | 0.26 |
| other liver disease | 1.14 (0.95-1.36) | 0.15 | 1.03 (0.82-1.29) | 0.81 | 1.26 (0.97-1.63) | 0.079 | 0.85 (0.25-2.87) | 0.79 |

**Table S15. Logistic Regression for All Factors and Outcomes for Patients Without Missing Data.** To identify whether BMI imputation could have introduced error, we repeated the analysis looking only at patients without data missingness. Odds ratios, 95% confidence intervals, and p-values for the association of features in all factors and outcomes.

| | Any Mental Illness | | Anxiety Disorder | | Mood Disorder | | Psychosis | |
|---|---|---|---|---|---|---|---|---|
| Predictor | OR (CI) | p-val | OR (CI) | p-val | OR (CI) | p-val | OR (CI) | p-val |
| (Intercept) | 0.04 (0.04-0.04) | <0.05 | 0.03 (0.03-0.03) | <0.05 | 0.02 (0.02-0.02) | <0.05 | 0 (0-0) | <0.05 |
| constitutional | 1.2 (1.14-1.26) | <0.05 | 1.25 (1.18-1.32) | <0.05 | 1.11 (1.02-1.19) | <0.05 | 1.17 (0.81-1.71) | 0.4 |
| neurological | 1.15 (1.1-1.2) | <0.05 | 1.11 (1.05-1.17) | <0.05 | 1.23 (1.16-1.32) | <0.05 | 1.45 (1.08-1.93) | <0.05 |
| gastrointestinal | 1.06 (1.01-1.11) | <0.05 | 1.09 (1.03-1.16) | <0.05 | 1.01 (0.94-1.09) | 0.714 | 1.36 (0.98-1.91) | 0.071 |
| pulmonary | 1 (0.94-1.06) | 0.923 | 0.98 (0.91-1.06) | 0.559 | 1.01 (0.92-1.11) | 0.858 | 1.01 (0.63-1.63) | 0.956 |
| cardiovascular | 1.34 (1.25-1.43) | <0.05 | 1.43 (1.32-1.55) | <0.05 | 1.07 (0.95-1.21) | 0.23 | 1.26 (0.74-2.15) | 0.398 |
| ENT | 1.01 (0.93-1.1) | 0.894 | 1.11 (1-1.22) | <0.05 | 0.87 (0.75-1.01) | 0.061 | 1 (0.48-2.07) | 0.995 |
| endocrine | 0.77 (0.69-0.86) | <0.05 | 0.79 (0.7-0.91) | <0.05 | 0.74 (0.62-0.89) | <0.05 | 0.3 (0.07-1.22) | 0.094 |
| skin | 0.73 (0.6-0.88) | <0.05 | 0.77 (0.62-0.96) | <0.05 | 0.62 (0.45-0.85) | <0.05 | null | null |
| laboratory | 0.99 (0.88-1.12) | 0.935 | 0.98 (0.84-1.14) | 0.797 | 1.04 (0.87-1.25) | 0.664 | null | null |
| eye | 0.61 (0.5-0.74) | <0.05 | 0.54 (0.42-0.7) | <0.05 | 0.74 (0.57-0.97) | <0.05 | null | null |
| immunology | 0.89 (0.61-1.28) | 0.521 | 0.91 (0.59-1.41) | 0.673 | 0.99 (0.57-1.71) | 0.963 | null | null |
| BMI under 20 | 0.44 (0.4-0.47) | <0.05 | 0.51 (0.46-0.56) | <0.05 | 0.27 (0.23-0.31) | <0.05 | 0.35 (0.13-0.93) | <0.05 |
| BMI 20 to 25 | 0.77 (0.73-0.82) | <0.05 | 0.8 (0.75-0.86) | <0.05 | 0.66 (0.61-0.73) | <0.05 | 1.43 (0.83-2.46) | 0.19 |
| BMI 25 to 30 | 0.82 (0.78-0.86) | <0.05 | 0.86 (0.81-0.92) | <0.05 | 0.7 (0.65-0.76) | <0.05 | 1.03 (0.61-1.73) | 0.922 |
| BMI 30 to 35 | 0.84 (0.79-0.88) | <0.05 | 0.87 (0.81-0.93) | <0.05 | 0.74 (0.68-0.8) | <0.05 | 1.19 (0.71-1.97) | 0.511 |
| BMI 35 to 40 | 0.89 (0.84-0.94) | <0.05 | 0.89 (0.83-0.95) | <0.05 | 0.86 (0.79-0.94) | <0.05 | 1.3 (0.75-2.24) | 0.348 |
| gender MALE | 0.56 (0.54-0.58) | <0.05 | 0.54 (0.52-0.56) | <0.05 | 0.54 (0.51-0.57) | <0.05 | 1.28 (0.96-1.72) | 0.1 |
| gender FEMALE | 1 | REF | 1 | REF | 1 | REF | 1 | REF |
| smoking status | 1.31 (1.18-1.46) | <0.05 | 1.28 (1.12-1.47) | <0.05 | 1.34 (1.14-1.56) | <0.05 | 2.53 (1.17-5.49) | <0.05 |
| race ethnicity White Non Hispanic | 1 | REF | 1 | REF | 1 | REF | 1 | REF |
| race ethnicity Other Non Hispanic | 0.74 (0.64-0.86) | <0.05 | 0.66 (0.54-0.79) | <0.05 | 0.82 (0.65-1.03) | 0.075 | 1.9 (0.7-5.17) | 0.207 |
| race ethnicity Native Hawaiian or Other Pacific Islander Non Hispanic | 0.36 (0.21-0.63) | <0.05 | 0.36 (0.19-0.69) | <0.05 | 0.33 (0.14-0.79) | <0.05 | null | null |
| race ethnicity Hispanic or Latino Any Race | 0.92 (0.88-0.97) | <0.05 | 0.89 (0.84-0.94) | <0.05 | 0.92 (0.86-0.99) | <0.05 | 1.04 (0.66-1.65) | 0.858 |
| race ethnicity Unknown | 0.73 (0.67-0.78) | <0.05 | 0.7 (0.64-0.77) | <0.05 | 0.73 (0.65-0.82) | <0.05 | 1.02 (0.53-1.96) | 0.943 |
| race ethnicity Black or African American Non Hispanic | 0.73 (0.7-0.77) | <0.05 | 0.65 (0.61-0.69) | <0.05 | 0.82 (0.76-0.88) | <0.05 | 1.63 (1.15-2.31) | <0.05 |
| race ethnicity Asian Non Hispanic | 0.58 (0.52-0.65) | <0.05 | 0.52 (0.45-0.6) | <0.05 | 0.61 (0.51-0.73) | <0.05 | 0.84 (0.31-2.28) | 0.732 |
| chronic resp | 1.12 (1.07-1.17) | <0.05 | 1.13 (1.07-1.19) | <0.05 | 1.07 (1-1.15) | <0.05 | 0.93 (0.63-1.38) | 0.738 |
| diabetes2 | 0.96 (0.91-1.01) | 0.202 | 0.89 (0.83-0.95) | <0.05 | 1.04 (0.96-1.13) | 0.293 | 0.9 (0.58-1.39) | 0.635 |
| hepatic steatosis | 1.03 (0.94-1.13) | 0.504 | 1.08 (0.96-1.22) | 0.164 | 1.01 (0.88-1.17) | 0.942 | null | null |
| hypertension | 0.97 (0.93-1.01) | 0.134 | 0.92 (0.88-0.97) | <0.05 | 0.95 (0.89-1.01) | 0.103 | 1.3 (0.91-1.86) | 0.16 |
| hypertensive kidney disease | 0.97 (0.85-1.1) | 0.619 | 1 (0.84-1.19) | 0.957 | 0.93 (0.78-1.11) | 0.414 | 0.45 (0.17-1.19) | 0.11 |
| ischemic heart disease | 0.94 (0.86-1.03) | 0.173 | 0.9 (0.8-1.01) | 0.06 | 1.06 (0.93-1.21) | 0.399 | 0.74 (0.38-1.44) | 0.379 |
| neoplasm | 0.93 (0.9-0.97) | <0.05 | 0.92 (0.88-0.97) | <0.05 | 0.88 (0.83-0.93) | <0.05 | 0.74 (0.51-1.07) | 0.115 |
| nicotine dependence | 1.23 (1.13-1.35) | <0.05 | 1.21 (1.08-1.35) | <0.05 | 1.35 (1.18-1.54) | <0.05 | 1.34 (0.66-2.73) | 0.428 |
| non hypertensive chronic kidney disease | 1.06 (0.97-1.16) | 0.198 | 0.9 (0.79-1.01) | 0.081 | 1.3 (1.14-1.47) | <0.05 | 1.49 (0.82-2.72) | 0.19 |
| non ischemic heart disease | 0.98 (0.93-1.03) | 0.482 | 0.93 (0.88-0.99) | <0.05 | 1.01 (0.94-1.09) | 0.871 | 1.57 (1.07-2.3) | <0.05 |
| other liver disease | 1.15 (1.05-1.26) | <0.05 | 1.07 (0.96-1.2) | 0.201 | 1.25 (1.1-1.42) | <0.05 | 1.57 (0.81-3.03) | 0.178 |

**Table S16. Logistic Regression for All Factors and Outcomes for 48-365 Day Early Post-Acute Phase Definition.** We performed a sensitivity analysis of the main finding by varying the definition for the early post-acute phase by beginning it at 48 days after diagnosis and ending at 365 days. Odds ratios, 95% confidence intervals, and p-values for the association of features in all factors and outcomes.

| | Any Mental Illness | | Anxiety Disorder | | Mood Disorder | | Psychosis | |
|---|---|---|---|---|---|---|---|---|
| Predictor | OR (CI) | p-val | OR (CI) | p-val | OR (CI) | p-val | OR (CI) | p-val |
| (Intercept) | 0.07 (0.07-0.07) | <0.05 | 0.05 (0.05-0.05) | <0.05 | 0.04 (0.03-0.04) | <0.05 | 0 (0-0) | <0.05 |
| constitutional | 1.12 (1.09-1.14) | <0.05 | 1.14 (1.11-1.17) | <0.05 | 1.05 (1.01-1.09) | <0.05 | 1.03 (0.85-1.26) | 0.75 |
| neurological | 1.13 (1.11-1.16) | <0.05 | 1.12 (1.09-1.15) | <0.05 | 1.13 (1.09-1.17) | <0.05 | 1.38 (1.18-1.61) | <0.05 |
| gastrointestinal | 1.08 (1.05-1.1) | <0.05 | 1.11 (1.08-1.14) | <0.05 | 1.05 (1.01-1.09) | <0.05 | 1.1 (0.91-1.33) | 0.33 |
| pulmonary | 0.9 (0.87-0.94) | <0.05 | 0.9 (0.87-0.94) | <0.05 | 0.91 (0.86-0.95) | <0.05 | 0.65 (0.47-0.91) | <0.05 |
| cardiovascular | 1.18 (1.13-1.23) | <0.05 | 1.25 (1.2-1.31) | <0.05 | 1.04 (0.98-1.11) | 0.2 | 1.39 (1.06-1.83) | <0.05 |
| ENT | 0.96 (0.92-1) | 0.063 | 0.99 (0.94-1.04) | 0.72 | 0.87 (0.82-0.93) | <0.05 | 0.67 (0.44-1.02) | 0.064 |
| endocrine | 0.85 (0.81-0.9) | <0.05 | 0.8 (0.75-0.85) | <0.05 | 0.96 (0.89-1.03) | 0.27 | 0.91 (0.59-1.4) | 0.68 |
| skin | 0.93 (0.85-1.01) | 0.073 | 0.94 (0.85-1.04) | 0.2 | 0.87 (0.77-0.99) | <0.05 | 0.94 (0.47-1.89) | 0.87 |
| laboratory | 0.86 (0.8-0.92) | <0.05 | 0.81 (0.74-0.88) | <0.05 | 0.93 (0.84-1.02) | 0.11 | 0.98 (0.6-1.58) | 0.92 |
| eye | 0.71 (0.65-0.78) | <0.05 | 0.64 (0.57-0.71) | <0.05 | 0.77 (0.69-0.87) | <0.05 | 0.62 (0.29-1.36) | 0.23 |
| BMI under 20 | 0.66 (0.6-0.72) | <0.05 | 0.71 (0.65-0.78) | <0.05 | 0.54 (0.47-0.61) | <0.05 | 0.41 (0.21-0.8) | <0.05 |
| BMI 20 to 25 | 0.89 (0.83-0.94) | <0.05 | 0.92 (0.85-0.99) | <0.05 | 0.82 (0.76-0.88) | <0.05 | 1.05 (0.68-1.62) | 0.83 |
| BMI 25 to 30 | 0.85 (0.8-0.9) | <0.05 | 0.87 (0.81-0.94) | <0.05 | 0.8 (0.74-0.86) | <0.05 | 1.15 (0.77-1.71) | 0.51 |
| BMI 30 to 35 | 0.88 (0.83-0.93) | <0.05 | 0.89 (0.83-0.95) | <0.05 | 0.84 (0.78-0.9) | <0.05 | 1.06 (0.72-1.56) | 0.77 |
| BMI 35 to 40 | 0.91 (0.84-0.99) | <0.05 | 0.92 (0.84-1.02) | 0.11 | 0.87 (0.8-0.96) | <0.05 | 0.98 (0.65-1.49) | 0.94 |
| gender MALE | 0.51 (0.5-0.52) | <0.05 | 0.49 (0.48-0.51) | <0.05 | 0.48 (0.46-0.49) | <0.05 | 1.02 (0.86-1.21) | 0.8 |
| gender FEMALE | 1 | REF | 1 | REF | 1 | REF | 1 | REF |
| smoking status | 1.09 (1-1.19) | 0.053 | 1.1 (0.99-1.22) | 0.08 | 1.05 (0.93-1.18) | 0.47 | 1.07 (0.57-2.01) | 0.84 |
| race ethnicity White Non Hispanic | 1 | REF | 1 | REF | 1 | REF | 1 | REF |
| race ethnicity Other Non Hispanic | 0.64 (0.58-0.7) | <0.05 | 0.61 (0.54-0.68) | <0.05 | 0.64 (0.56-0.73) | <0.05 | 1.31 (0.7-2.47) | 0.4 |
| race ethnicity Native Hawaiian or Other Pacific Islander Non Hispanic | 0.67 (0.48-0.93) | <0.05 | 0.65 (0.43-0.97) | <0.05 | 0.76 (0.49-1.18) | 0.22 | null | null |
| race ethnicity Hispanic or Latino Any Race | 1.04 (1.01-1.07) | <0.05 | 1.01 (0.97-1.05) | 0.56 | 1.04 (0.99-1.08) | 0.1 | 1.16 (0.89-1.52) | 0.27 |
| race ethnicity Unknown | 0.78 (0.73-0.82) | <0.05 | 0.74 (0.69-0.79) | <0.05 | 0.77 (0.7-0.83) | <0.05 | 0.79 (0.46-1.36) | 0.4 |
| race ethnicity Black or African American Non Hispanic | 0.85 (0.83-0.88) | <0.05 | 0.78 (0.75-0.81) | <0.05 | 0.88 (0.84-0.92) | <0.05 | 2.08 (1.69-2.56) | <0.05 |
| race ethnicity Asian Non Hispanic | 0.63 (0.58-0.68) | <0.05 | 0.62 (0.56-0.68) | <0.05 | 0.56 (0.49-0.63) | <0.05 | 1.11 (0.64-1.94) | 0.71 |
| chronic resp | 1.36 (1.32-1.41) | <0.05 | 1.42 (1.37-1.47) | <0.05 | 1.27 (1.21-1.33) | <0.05 | 0.99 (0.74-1.32) | 0.93 |
| diabetes2 | 1.02 (0.98-1.05) | 0.41 | 0.89 (0.85-0.93) | <0.05 | 1.17 (1.11-1.23) | <0.05 | 0.99 (0.74-1.31) | 0.92 |
| hepatic steatosis | 1.15 (1.07-1.23) | <0.05 | 1.14 (1.05-1.24) | <0.05 | 1.15 (1.04-1.27) | <0.05 | 0.45 (0.19-1.02) | 0.057 |
| hypertension | 1.12 (1.09-1.15) | <0.05 | 1.07 (1.03-1.11) | <0.05 | 1.12 (1.08-1.17) | <0.05 | 1.17 (0.93-1.47) | 0.19 |
| hypertensive kidney disease | 1.03 (0.94-1.14) | 0.49 | 1.06 (0.93-1.2) | 0.39 | 1.03 (0.9-1.16) | 0.69 | 0.5 (0.25-1.02) | 0.058 |
| ischemic heart disease | 0.96 (0.9-1.03) | 0.25 | 0.88 (0.81-0.96) | <0.05 | 1.06 (0.97-1.16) | 0.16 | 0.87 (0.55-1.39) | 0.57 |
| neoplasm | 1.03 (1-1.06) | 0.087 | 1.03 (1-1.07) | 0.076 | 0.95 (0.92-1) | <0.05 | 0.54 (0.4-0.72) | <0.05 |
| nicotine dependence | 1.54 (1.44-1.64) | <0.05 | 1.56 (1.44-1.69) | <0.05 | 1.6 (1.46-1.75) | <0.05 | 1.61 (0.99-2.61) | 0.053 |
| non hypertensive chronic kidney disease | 0.91 (0.85-0.97) | <0.05 | 0.8 (0.73-0.87) | <0.05 | 1.03 (0.94-1.13) | 0.48 | 1.3 (0.85-1.99) | 0.23 |
| non ischemic heart disease | 0.98 (0.95-1.02) | 0.36 | 0.94 (0.9-0.98) | <0.05 | 1.02 (0.97-1.07) | 0.51 | 1.54 (1.18-2) | <0.05 |
| other liver disease | 1.13 (1.06-1.21) | <0.05 | 1.15 (1.06-1.24) | <0.05 | 1.11 (1.01-1.22) | <0.05 | 1.77 (1.1-2.84) | <0.05 |

**Table S17. Logistic Regression for All Factors and Outcomes Excluding PASC-AMs within 10 Days of Psychiatric Disease.** We performed a sensitivity analysis of the main finding by excluding PASC-AMs within 10 days of psychiatric disease. Odds ratios, 95% confidence intervals, and p-values for the association of features in all factors and outcomes.

| | Any Mental Illness | | Anxiety Disorder | | Mood Disorder | | Psychosis | |
|---|---|---|---|---|---|---|---|---|
| Predictor | OR (CI) | p-val | OR (CI) | p-val | OR (CI) | p-val | OR (CI) | p-val |
| (Intercept) | 0.02 (0.02-0.03) | <0.05 | 0.02 (0.02-0.02) | <0.05 | 0.01 (0.01-0.01) | <0.05 | 0 (0-0) | <0.05 |
| constitutional | 1.06 (1.01-1.11) | <0.05 | 1.1 (1.04-1.16) | <0.05 | 1.02 (0.95-1.09) | 0.56 | 0.91 (0.62-1.33) | 0.62 |
| neurological | 1.04 (0.99-1.09) | 0.09 | 1.02 (0.96-1.07) | 0.59 | 1.07 (1-1.14) | 0.054 | 1.32 (1-1.75) | 0.053 |
| gastrointestinal | 1 (0.95-1.04) | 0.85 | 1.03 (0.97-1.08) | 0.33 | 0.95 (0.88-1.02) | 0.14 | 1.09 (0.77-1.54) | 0.62 |
| pulmonary | 0.93 (0.88-0.99) | <0.05 | 0.91 (0.85-0.98) | <0.05 | 0.95 (0.87-1.04) | 0.28 | 0.94 (0.6-1.48) | 0.79 |
| cardiovascular | 1.16 (1.08-1.24) | <0.05 | 1.23 (1.13-1.33) | <0.05 | 0.96 (0.86-1.08) | 0.48 | 1.25 (0.76-2.04) | 0.38 |
| ENT | 0.91 (0.84-0.99) | <0.05 | 0.97 (0.88-1.07) | 0.51 | 0.85 (0.74-0.97) | <0.05 | 0.81 (0.39-1.68) | 0.58 |
| endocrine | 0.78 (0.7-0.86) | <0.05 | 0.8 (0.71-0.91) | <0.05 | 0.78 (0.67-0.91) | <0.05 | 0.25 (0.06-0.99) | <0.05 |
| BMI under 20 | 0.57 (0.53-0.62) | <0.05 | 0.62 (0.57-0.68) | <0.05 | 0.45 (0.4-0.5) | <0.05 | 0.4 (0.21-0.76) | <0.05 |
| BMI 20 to 25 | 0.85 (0.79-0.92) | <0.05 | 0.87 (0.8-0.95) | <0.05 | 0.78 (0.72-0.85) | <0.05 | 1.06 (0.71-1.57) | 0.78 |
| BMI 25 to 30 | 0.85 (0.8-0.91) | <0.05 | 0.87 (0.8-0.94) | <0.05 | 0.8 (0.74-0.87) | <0.05 | 0.99 (0.63-1.55) | 0.96 |
| BMI 30 to 35 | 0.88 (0.82-0.94) | <0.05 | 0.89 (0.83-0.96) | <0.05 | 0.83 (0.77-0.9) | <0.05 | 1.12 (0.74-1.7) | 0.6 |
| BMI 35 to 40 | 0.91 (0.83-1) | <0.05 | 0.91 (0.82-1.01) | 0.072 | 0.89 (0.81-0.99) | <0.05 | 1 (0.59-1.7) | 1 |
| gender MALE | 0.54 (0.52-0.55) | <0.05 | 0.52 (0.5-0.53) | <0.05 | 0.51 (0.49-0.53) | <0.05 | 1.12 (0.92-1.36) | 0.27 |
| gender FEMALE | 1 | REF | 1 | REF | 1 | REF | 1 | REF |
| smoking status | 1.35 (1.24-1.48) | <0.05 | 1.34 (1.21-1.49) | <0.05 | 1.34 (1.18-1.52) | <0.05 | 1.66 (0.93-2.93) | 0.084 |
| race ethnicity White Non Hispanic | 1 | REF | 1 | REF | 1 | REF | 1 | REF |
| race ethnicity Other Non Hispanic | 0.66 (0.59-0.73) | <0.05 | 0.63 (0.55-0.72) | <0.05 | 0.66 (0.55-0.77) | <0.05 | 1.07 (0.48-2.42) | 0.86 |
| race ethnicity Native Hawaiian or Other Pacific Islander Non Hispanic | 0.68 (0.46-1.02) | 0.06 | 0.63 (0.39-1.04) | 0.069 | 0.72 (0.41-1.28) | 0.26 | null | null |
| race ethnicity Hispanic or Latino Any Race | 1.04 (1-1.08) | <0.05 | 1.01 (0.97-1.05) | 0.69 | 1.01 (0.96-1.07) | 0.6 | 1.26 (0.92-1.73) | 0.15 |
| race ethnicity Unknown | 0.87 (0.81-0.92) | <0.05 | 0.87 (0.81-0.94) | <0.05 | 0.83 (0.75-0.91) | <0.05 | 1.11 (0.66-1.87) | 0.7 |
| race ethnicity Black or African American Non Hispanic | 0.84 (0.81-0.87) | <0.05 | 0.76 (0.73-0.8) | <0.05 | 0.88 (0.83-0.93) | <0.05 | 2.36 (1.88-2.97) | <0.05 |
| race ethnicity Asian Non Hispanic | 0.69 (0.63-0.76) | <0.05 | 0.65 (0.59-0.73) | <0.05 | 0.65 (0.57-0.75) | <0.05 | 1.13 (0.58-2.2) | 0.73 |
| chronic resp | 1.33 (1.28-1.38) | <0.05 | 1.37 (1.31-1.43) | <0.05 | 1.23 (1.16-1.3) | <0.05 | 1.02 (0.75-1.38) | 0.92 |
| diabetes2 | 1.01 (0.97-1.06) | 0.53 | 0.93 (0.88-0.98) | <0.05 | 1.13 (1.06-1.2) | <0.05 | 0.98 (0.71-1.37) | 0.93 |
| hepatic steatosis | 1.13 (1.04-1.22) | <0.05 | 1.17 (1.06-1.29) | <0.05 | 1.12 (1-1.26) | 0.056 | 0.17 (0.04-0.68) | <0.05 |
| hypertension | 1.14 (1.1-1.18) | <0.05 | 1.08 (1.04-1.13) | <0.05 | 1.13 (1.08-1.19) | <0.05 | 1.01 (0.77-1.32) | 0.96 |
| hypertensive kidney disease | 1.06 (0.95-1.18) | 0.29 | 1.08 (0.94-1.24) | 0.29 | 1.02 (0.89-1.18) | 0.75 | 0.94 (0.47-1.88) | 0.86 |
| ischemic heart disease | 0.98 (0.91-1.05) | 0.5 | 0.92 (0.84-1.01) | 0.088 | 1.1 (0.99-1.21) | 0.078 | 0.89 (0.54-1.46) | 0.64 |
| neoplasm | 1.11 (1.07-1.15) | <0.05 | 1.12 (1.08-1.16) | <0.05 | 1.02 (0.97-1.07) | 0.44 | 0.75 (0.56-1.01) | 0.058 |
| nicotine dependence | 1.41 (1.32-1.52) | <0.05 | 1.42 (1.31-1.54) | <0.05 | 1.51 (1.37-1.67) | <0.05 | 1.76 (1.09-2.85) | <0.05 |
| non hypertensive chronic kidney disease | 1.01 (0.93-1.09) | 0.89 | 0.88 (0.79-0.97) | <0.05 | 1.17 (1.05-1.3) | <0.05 | 1.23 (0.74-2.05) | 0.43 |
| non ischemic heart disease | 1.07 (1.03-1.12) | <0.05 | 1.03 (0.98-1.09) | 0.22 | 1.07 (1.01-1.14) | <0.05 | 1.76 (1.31-2.35) | <0.05 |
| other liver disease | 1.25 (1.16-1.34) | <0.05 | 1.21 (1.1-1.32) | <0.05 | 1.28 (1.15-1.43) | <0.05 | 1.97 (1.22-3.2) | <0.05 |

**Table S18. Chi-Square Analysis of Differential Rates of HPO Terms Between Outcomes:** In this analysis we perform a chi-square test to determine whether individual HPO terms are present at different rates in the groups of patients with each outcome. Count gives the total number of patients with the given term. P-val provides the Bonferroni corrected p-values for whether the difference between outcome groups is significant. Category provides the PASC-AM category that the HPO term was included in for the main analysis.

| HPO Term | count | p-val (adj) | category |
|---|---|---|---|
| Chest pain (HP:0100749) | 27266 | $1.86 \times 10^{-121}$ | constitutional |
| Tachycardia (HP:0001649) | 8666 | $9.90 \times 10^{-84}$ | cardiovascular |
| Confusion (HP:0001289) | 1451 | $2.97 \times 10^{-67}$ | neurological |
| Palpitations (HP:0001962) | 10316 | $9.24 \times 10^{-61}$ | cardiovascular |
| Asthenia (HP:0025406) | 6907 | $1.35 \times 10^{-52}$ | constitutional |
| Fatigue (HP:0012378) | 22318 | $5.42 \times 10^{-48}$ | constitutional |
| Vertigo (HP:0002321) | 11814 | $4.60 \times 10^{-46}$ | ENT |
| Encephalopathy (HP:0001298) | 2518 | $4.01 \times 10^{-44}$ | neurological |
| Hypotension (HP:0002615) | 3023 | $6.83 \times 10^{-33}$ | cardiovascular |
| Parkinsonism (HP:0001300) | $< 20$ | $2.78 \times 10^{-30}$ | neurological |
| Hypoxemia (HP:0012418) | 5963 | $1.57 \times 10^{-28}$ | pulmonary |
| Headache (HP:0002315) | 14406 | $1.42 \times 10^{-27}$ | neurological |
| Insomnia (HP:0100785) | 6050 | $7.23 \times 10^{-25}$ | neurological |
| Nausea (HP:0002018) | 7559 | $1.40 \times 10^{-24}$ | gastrointestinal |
| Abdominal pain (HP:0002027) | 17339 | $6.42 \times 10^{-24}$ | gastrointestinal |
| Atelectasis (HP:0100750) | 2708 | $1.27 \times 10^{-18}$ | pulmonary |
| Dysphagia (HP:0002015) | 6380 | $4.52 \times 10^{-18}$ | neurological |
| Diarrhea (HP:0002014) | 10879 | $5.83 \times 10^{-18}$ | gastrointestinal |
| Seizure (HP:0001250) | 5898 | $5.19 \times 10^{-16}$ | neurological |
| Constipation (HP:0002019) | 13037 | $5.61 \times 10^{-16}$ | gastrointestinal |
| Skeletal muscle atrophy (HP:0003202) | 356 | $3.41 \times 10^{-12}$ | neurological |
| Pulmonary embolism (HP:0002204) | 4883 | $8.28 \times 10^{-10}$ | pulmonary |
| Tremor (HP:0001337) | 1596 | $2.27 \times 10^{-9}$ | neurological |
| Stroke (HP:0001297) | 110 | $2.94 \times 10^{-9}$ | cardiovascular |
| Syncope (HP:0001279) | 848 | $1.02 \times 10^{-8}$ | cardiovascular |
| Memory impairment (HP:0002354) | 2367 | $3.51 \times 10^{-8}$ | neurological |
| Gastroparesis (HP:0002578) | 686 | $7.98 \times 10^{-8}$ | gastrointestinal |
| Paresthesia (HP:0003401) | 4761 | $4.14 \times 10^{-7}$ | neurological |
| Slurred speech (HP:0001350) | 234 | $5.63 \times 10^{-7}$ | neurological |
| Myalgia (HP:0003326) | 5446 | $1.45 \times 10^{-6}$ | constitutional |
| Migraine (HP:0002076) | 4576 | $3.39 \times 10^{-6}$ | neurological |
| Bronchiectasis (HP:0002110) | 1074 | $1.52 \times 10^{-5}$ | pulmonary |
| Polyneuropathy (HP:0001271) | 4504 | $5.08 \times 10^{-5}$ | neurological |
| Shivering (HP:0025144) | 1028 | $9.04 \times 10^{-5}$ | constitutional |
| Limb pain (HP:0009763) | 3048 | $1.99 \times 10^{-4}$ | constitutional |
| Thrombocytopenia (HP:0001873) | 4806 | $2.55 \times 10^{-4}$ | laboratory |
| Aphasia (HP:0002381) | 495 | $5.63 \times 10^{-4}$ | neurological |
| Dyspnea (HP:0002094) | 3778 | $5.92 \times 10^{-4}$ | pulmonary |
| Hepatic steatosis (HP:0001397) | 5735 | $7.51 \times 10^{-4}$ | gastrointestinal |
| Bradycardia (HP:0001662) | 4288 | $1.78 \times 10^{-3}$ | cardiovascular |
| Pulmonary fibrosis (HP:0002206) | 1321 | $3.92 \times 10^{-3}$ | pulmonary |

Continued on next page

**Table S18 – continued from previous page**

| HPO Term | count | p-val (adj) | category |
|---|---|---|---|
| Malnutrition (HP:0004395) | 507 | $3.36 \times 10^{-2}$ | gastrointestinal |
| Hyperglycemia (HP:0003074) | 6760 | $6.92 \times 10^{-2}$ | laboratory |
| Tinnitus (HP:0000360) | 1450 | $7.44 \times 10^{-2}$ | ENT |
| Hyperkinetic movements (HP:0002487) | 4140 | $1.06 \times 10^{-1}$ | neurological |
| Hepatitis (HP:0012115) | 236 | $1.07 \times 10^{-1}$ | gastrointestinal |
| Hepatomegaly (HP:0002240) | 938 | $1.17 \times 10^{-1}$ | gastrointestinal |
| Hypoglycemia (HP:0001943) | 1257 | $1.19 \times 10^{-1}$ | laboratory |
| Muscle weakness (HP:0001324) | 2756 | $1.24 \times 10^{-1}$ | neurological |
| Pain (HP:0012531) | 5019 | $1.32 \times 10^{-1}$ | constitutional |
| Cough (HP:0012735) | 26334 | $2.12 \times 10^{-1}$ | pulmonary |
| Dysarthria (HP:0001260) | 372 | $3.61 \times 10^{-1}$ | neurological |
| Orthostatic hypotension (HP:0001278) | 1156 | $4.71 \times 10^{-1}$ | neurological |
| Exercise intolerance (HP:0003546) | $< 20$ | $7.37 \times 10^{-1}$ | constitutional |
| Abnormal reflex (HP:0031826) | 141 | 1.0 | neurological |
| Abnormality of movement (HP:0100022) | 155 | 1.0 | neurological |
| Ageusia (HP:0041051) | $< 20$ | 1.0 | neurological |
| Agnosia (HP:0010524) | 48 | 1.0 | neurological |
| Alopecia (HP:0001596) | 126 | 1.0 | skin |
| Anaphylactic shock (HP:0100845) | 246 | 1.0 | immunology |
| Angina pectoris (HP:0001681) | 1965 | 1.0 | cardiovascular |
| Anosmia (HP:0000458) | 694 | 1.0 | neurological |
| Antinuclear antibody positivity (HP:0003493) | 60 | 1.0 | immunology |
| Arthralgia (HP:0002829) | 3690 | 1.0 | constitutional |
| Arthritis (HP:0001369) | 192 | 1.0 | constitutional |
| Ataxia (HP:0001251) | 336 | 1.0 | neurological |
| Babinski sign (HP:0003487) | $< 20$ | 1.0 | neurological |
| Blindness (HP:0000618) | $< 20$ | 1.0 | eye |
| Blurred vision (HP:0000622) | 47 | 1.0 | eye |
| Bone pain (HP:0002653) | $< 20$ | 1.0 | constitutional |
| Bowel incontinence (HP:0002607) | 734 | 1.0 | gastrointestinal |
| Bradykinesia (HP:0002067) | $< 20$ | 1.0 | neurological |
| Chest tightness (HP:0031352) | 47 | 1.0 | constitutional |
| Conjunctivitis (HP:0000509) | 955 | 1.0 | eye |
| Dermatographic urticaria (HP:0011971) | 148 | 1.0 | skin |
| Diabetes mellitus (HP:0000819) | 509 | 1.0 | endocrine |
| Difficulty walking (HP:0002355) | 1321 | 1.0 | constitutional |
| Diplopia (HP:0000651) | 538 | 1.0 | eye |
| Dysmetria (HP:0001310) | $< 20$ | 1.0 | neurological |
| Dystonia (HP:0001332) | 241 | 1.0 | neurological |
| Ear pain (HP:0030766) | 3251 | 1.0 | ENT |
| Edema (HP:0000969) | 4358 | 1.0 | endocrine |
| Elevated circulating alkaline phosphatase concentration (HP:0003155) | $< 20$ | 1.0 | laboratory |
| Elevated circulating C reactive protein concentration (HP:0011227) | 90 | 1.0 | laboratory |
| Elevated circulating creatinine concentration (HP:0003259) | 51 | 1.0 | laboratory |

Continued on next page

**Table S18 – continued from previous page**

| HPO Term | count | p-val (adj) | category |
|---|---|---|---|
| Elevated circulating thyroid stimulating hormone concentration (HP:0002925) | 34 | 1.0 | laboratory |
| Elevated erythrocyte sedimentation rate (HP:0003565) | 456 | 1.0 | laboratory |
| Exertional dyspnea (HP:0002875) | 510 | 1.0 | pulmonary |
| Expressive aphasia (HP:0002427) | < 20 | 1.0 | neurological |
| Fever (HP:0001945) | 14170 | 1.0 | endocrine |
| Gastric ulcer (HP:0002592) | < 20 | 1.0 | gastrointestinal |
| Gastroesophageal reflux (HP:0002020) | 5106 | 1.0 | gastrointestinal |
| Hand muscle weakness (HP:0030237) | < 20 | 1.0 | neurological |
| Hearing impairment (HP:0000365) | 2466 | 1.0 | ENT |
| Heat intolerance (HP:0002046) | < 20 | 1.0 | endocrine |
| Hemoptysis (HP:0002105) | 675 | 1.0 | pulmonary |
| Hyperacusis (HP:0010780) | 48 | 1.0 | ENT |
| Hyperesthesia (HP:0100963) | 33 | 1.0 | neurological |
| Hyperhidrosis (HP:0000975) | 22 | 1.0 | skin |
| Hypertension (HP:0000822) | 663 | 1.0 | cardiovascular |
| Hypocalcemia (HP:0002901) | 1178 | 1.0 | laboratory |
| Hypophosphatemia (HP:0002148) | < 20 | 1.0 | laboratory |
| Hyposmia (HP:0004409) | < 20 | 1.0 | neurological |
| Hypothermia (HP:0002045) | 118 | 1.0 | endocrine |
| Impaired executive functioning (HP:0033051) | < 20 | 1.0 | neurological |
| Irregular menstruation (HP:0000858) | 3587 | 1.0 | endocrine |
| Keratoconjunctivitis sicca (HP:0001097) | 5122 | 1.0 | eye |
| Low grade fever (HP:0011134) | < 20 | 1.0 | endocrine |
| Lymphadenopathy (HP:0002716) | 1185 | 1.0 | immunology |
| Lymphopenia (HP:0001888) | 309 | 1.0 | immunology |
| Male sexual dysfunction (HP:0040307) | < 20 | 1.0 | endocrine |
| Menorrhagia (HP:0000132) | 364 | 1.0 | endocrine |
| Muscle spasm (HP:0003394) | 67 | 1.0 | neurological |
| Myocarditis (HP:0012819) | 191 | 1.0 | cardiovascular |
| Nasal congestion (HP:0001742) | 7667 | 1.0 | ENT |
| Night sweats (HP:0030166) | < 20 | 1.0 | constitutional |
| Nonproductive cough (HP:0031246) | < 20 | 1.0 | pulmonary |
| Ocular pain (HP:0200026) | 881 | 1.0 | eye |
| Pancreatitis (HP:0001733) | < 20 | 1.0 | endocrine |
| Parageusia (HP:0031249) | 565 | 1.0 | neurological |
| Pericardial effusion (HP:0001698) | 26 | 1.0 | cardiovascular |
| Petechiae (HP:0000967) | < 20 | 1.0 | skin |
| Pharyngalgia (HP:0033050) | 1208 | 1.0 | ENT |
| Photophobia (HP:0000613) | < 20 | 1.0 | eye |
| Pleural thickening (HP:0031944) | < 20 | 1.0 | pulmonary |
| Pleuritis (HP:0002102) | 665 | 1.0 | pulmonary |
| Productive cough (HP:0031245) | < 20 | 1.0 | pulmonary |
| Pruritus (HP:0000989) | 447 | 1.0 | skin |
| Pulsatile tinnitus (HP:0008629) | 183 | 1.0 | ENT |
| Recurrent fever (HP:0001954) | 71 | 1.0 | endocrine |

Continued on next page

**Table S18 – continued from previous page**

| HPO Term | count | p-val (adj) | category |
|---|---|---|---|
| Red eye (HP:0025337) | < 20 | 1.0 | eye |
| Renal insufficiency (HP:0000083) | 416 | 1.0 | endocrine |
| Restless legs (HP:0012452) | 1928 | 1.0 | neurological |
| Restrictive ventilatory defect (HP:0002091) | 32 | 1.0 | pulmonary |
| Rhinitis (HP:0012384) | 30 | 1.0 | ENT |
| Rhinorrhea (HP:0031417) | 74 | 1.0 | pulmonary |
| Rhonchi (HP:0030831) | < 20 | 1.0 | pulmonary |
| Scaling skin (HP:0040189) | < 20 | 1.0 | skin |
| Skin rash (HP:0000988) | 7835 | 1.0 | skin |
| Sleep apnea (HP:0010535) | 5030 | 1.0 | neurological |
| Sleep disturbance (HP:0002360) | 293 | 1.0 | neurological |
| Sneeze (HP:0025095) | 189 | 1.0 | pulmonary |
| Somatic sensory dysfunction (HP:0003474) | < 20 | 1.0 | neurological |
| Spasticity (HP:0001257) | 56 | 1.0 | neurological |
| Splenomegaly (HP:0001744) | 850 | 1.0 | gastrointestinal |
| Urinary incontinence (HP:0000020) | 2102 | 1.0 | endocrine |
| Venous thrombosis (HP:0004936) | 110 | 1.0 | cardiovascular |
| Vitreous floaters (HP:0100832) | 943 | 1.0 | eye |
| Vomiting (HP:0002013) | 4582 | 1.0 | gastrointestinal |
| Wheezing (HP:0030828) | 3320 | 1.0 | pulmonary |
| Xerostomia (HP:0000217) | 336 | 1.0 | constitutional |
